# Multi-Omics Modelling of Plasma pTau181, GFAP, and Metabolic Features Enables Risk Stratification in Prodromal Alzheimer’s Disease

**DOI:** 10.1101/2025.11.12.25339932

**Authors:** Tereza Kacerova, Jack J.J.J. Miller, Daniel E. Radford-Smith, Stephen M. Smith, Alexander F. Jeans, Fredrik Jernerén, Dawn Shepherd, Elisabete Pires, Sebastian de Jel, Qingxia Gong, Eric Schiffer, Laia Montoliu-Gaya, Nicholas J. Ashton, Thomas Olsen, David Leppert, Henrik Zetterberg, Helga Refsum, James S. O. McCullagh, A. David Smith, Daniel C. Anthony

**Author notes:** **Corresponding author:** Tereza Kacerova, Chemistry Research Laboratory, Department of Chemistry, University of Oxford, Oxford OX1 3TA, UK Kavli Institute of Nanoscience Discovery, Dorothy Crowfoot Hodgkin Building, Oxford OX1 3QU, UK Physical and Theoretical Chemistry, University of Oxford, Oxford OX1 3QZ, UK.

## Abstract

**Background:** Mild cognitive impairment (MCI) is a heterogeneous state between normal ageing and dementia, often considered prodromal to Alzheimer’s disease (AD). Progression is variable, and distinguishing stable from progressive MCI remains difficult, particularly in the presence of mixed neuropathology. Blood biomarkers such as phosphorylated tau181 (pTau181), glial fibrillary acidic protein (GFAP), and neurofilament light chain (NfL) demonstrate prognostic value in established AD, but limited performance for prognosticating progression from MCI.

**Methods:** Blood protein biomarkers (pTau181, GFAP, NfL) were integrated with NMR- and LC-MS-derived metabolomic features. In a deeply phenotyped MCI cohort (VITACOG; *n*=68) with two-year MRI follow-up, cross-validated logistic regression identified discriminative multi-analyte panels to distinguish stable from progressive MCI. Disease progression was defined by worsening cortical atrophy, measured via annualised brain volume loss. Generalisability was tested in a larger community-based cohort from UK Biobank (*n*=223) and two Oxford Project to Investigate Memory and Ageing (OPTIMA) subsets with histopathological diagnosis (*n*=61, *n*=37).

**Results:** Integration of pTau181 with six metabolite features yielded the highest prognostic performance (AUC 0.91; accuracy 80%), with metabolomic findings independently validated in the OPTIMA cohort. A complementary GFAP-NMR panel also performed strongly (AUC 0.80; accuracy 75%). In contrast, individual metabolites, including the atrophy marker homocysteine, and standalone protein biomarkers performed poorly (AUC ≤0.66), as well their combination (AUC 0.68), highlighting the added value of multi-omic integration. In an asymptomatic ageing population (UK Biobank), the models served as a population-level stress test, confirming that multi-omic integration improved specificity for MRI-derived atrophy measures and captured atrophy-related risk in community cohorts.

**Conclusion:** Multi-omic integration of protein and metabolic features markedly improved prognostication of MCI progression by capturing early neurodegenerative signatures, yielding translational panels suitable for scalable risk stratification and early therapeutic intervention in clinical practice.

## Background

Mild cognitive impairment (MCI) represents a clinically heterogeneous stage within the Alzheimer’s disease (AD) continuum, with trajectories shaped by mixed underlying pathologies that may lead to either long-term stability or progression^1^. Accurate identification of individuals at highest risk of conversion to overt dementia remains a major unmet need, as disease-modifying therapies are likely to be most effective when initiated during the early phases of the disease^2^. Neuropsychological testing lacks sufficient precision for individual-level prognostication, creating demand for biomarkers that detect individuals at risk of accelerated neurodegeneration. Cerebrospinal fluid (CSF) (tau/Aβ₄₂ ratios^3^) and imaging measures (hippocampal atrophy)^4,5^ prognosticate progression with high accuracy but are invasive, costly, and unsuitable for large-scale deployment. Moreover, these measures do not reliably distinguish AD from other dementias with overlapping or mixed pathologies. With disease-modifying therapies entering clinical use, scalable blood-based biomarkers are urgently needed to enable early, personalised intervention.

Among blood-based biomarkers, amyloid-β peptides, phosphorylated tau species (including pTau181 and pTau217)^6–9^, neurofilament light chain (NfL), and glial fibrillary acidic protein (GFAP)^10,11^ exhibit strong associations with AD pathology and cognitive decline. Combinations such as plasma pTau181 and NfL can identify amyloid- and tau-positive individuals (A+/T+)^9^. Recent regulatory approvals, including plasma-based assays such as the Roche Elecsys pTau181 and Lumipulse G pTau217/Aβ₄₂ ratio, underscore the clinical momentum of blood-based biomarkers^12,13^. Yet, their diagnostic accuracy as standalone markers remains limited in prognosticating the transition from MCI to AD and other types of dementing illnesses, underscoring the multifactorial pathogenesis of MCI^14^. Even within the tau pathway, MTBR-tau243 predominantly reflects tau-related pathology and cognitive decline^15^, illustrating that such markers capture only one dimension of AD-related biology rather than its full pathological spectrum.

Growing interest in multi-analyte models reflects the need to pair protein biomarkers with complementary molecular signatures that capture prognostic information across mixed pathologies, rather than reporting on single disease entities alone. Metabolomics provides a window into early biochemical disturbances in MCI, such as lipid dysregulation, oxidative stress, and altered energy metabolism, that are readily detectable in blood and reflect dynamic interactions among the underlying pathologies commonly present in MCI^16,17^. Plasma concentration of total homocysteine (tHcy), for example, has been associated with brain atrophy and neurodegeneration via impaired methylation, vascular injury, and neuronal excitotoxicity, but lacks specificity as an individual marker^18–20^. Integrating metabolomic measures with established proteins may therefore enhance prognostic accuracy while providing deeper mechanistic insight^16,17^.

Building on prior work in multiple sclerosis, where nuclear magnetic resonance (NMR)-based metabolomics integrated with GFAP and NfL enhanced prognostic models^18,19^, we applied a similar multimodal framework to a deeply phenotyped MCI cohort^21,22^. We hypothesised that compact, blood-based multi-analyte panels combining metabolic and protein markers would outperform individual biomarkers in prognosticating disease progression. The study further incorporated *post-mortem*-confirmed dementia cases to validate the metabolomic component and assess cross-stage reproducibility, and leveraged the UK Biobank as a large-scale test of whether molecular signatures of neurodegeneration identified in MCI extend to the general population. Together, these analyses aimed to establish a scalable, biologically grounded framework for blood-based risk stratification and early intervention in prodromal AD.

## Methods

### Cohort selection and validation design

The VITACOG trial (2004-2006, Oxford) was a randomised controlled study investigating the effects of B vitamin supplementation in older adults (>70 years) with amnestic or non-amnestic MCI, defined by Petersen’s criteria (1.0-1.5 standard deviation (SD) below normal)^19,23^. For this analysis, only baseline samples from the placebo arm were included to eliminate treatment-related confounding. Participants were classified as progressors or non-progressors according to whole-brain MRI atrophy rates over two years, using an annualised threshold of ≥0.72% to define progression, representing the upper limit of normal ageing and previously validated for differentiating pathological from age-related decline^24,25^. Independent validation was conducted in two subsets of the Oxford Project to Investigate Memory and Ageing (OPTIMA) - one with pure AD and one with mixed neuropathological diagnoses^26,27^. All selected OPTIMA individuals had neuropathologically confirmed dementia, but exhibited a Mini-Mental State Examination (MMSE) score ≥24 at the time of sampling. Stable cases (*n*=18 pure AD; *n*=35 mixed pathology) showed minimal cognitive change (ΔMMSE 0-1) over two years, whereas progressors (*n*=19 pure AD; *n*=26 mixed pathology) exhibited marked decline (ΔMMSE 4-13). Full cohort details are provided in the Supplementary Methods (eMethods 1, 2) and SI Tables 1, 2.

**Table 1.** Demographic and clinical characteristics of mild cognitive impairment (MCI) participants classified as stable or progressors based on annualised brain atrophy rates. Data are presented as mean ± standard deviation. Atrophy rates were derived from SIENA-based quantification of longitudinal whole-brain volume loss. Participants were classified as *progressors* if their annualised atrophy rate was ≥0.72%, and as *stable* if <0.72%. Between-group comparisons were performed using unpaired two-tailed t-tests for continuous variables and chi-square tests for categorical variables. A *p*-value < 0.05 was considered statistically significant.

| Characteristics | Stable<br>$n = 26$ | Progressor<br>$n = 42$ | $p$-value |
| --- | --- | --- | --- |
| Age (baseline) | 75.7 ± 3.6 | 77.4 ± 4.9 | 0.146 |
| Sex (% female) | 13 (56%) | 30 (71%) | 0.120 |
| Years of education | 15.0 ± 3.2 | 14.6 ± 3.3 | 0.642 |
| Body Mass Index, kg/m<sup>2</sup> (baseline) | 26.3 ± 4.2 | 26.9 ± 4.1 | 0.610 |
| Systolic blood pressure, mmHg (baseline) | 147 ± 15 | 147 ± 20 | 0.958 |
| Diastolic blood pressure, mmHg (baseline) | 82 ± 12 | 77 ± 12 | 0.056 |
| TICS-M (baseline) | 24.65 ± 2.73 | 25.12 ± 2.60 | 0.491 |
| TICS-M (follow-up) | 27.12 ± 3.41 | 25.31 ± 4.70 | 0.098 |
| MMSE (baseline) | 28.46 ± 1.15 | 28.19 ± 1.55 | 0.450 |
| MMSE (follow-up) | 27.77 ± 2.19 | 27.32 ± 2.70 | 0.482 |
| Brain volume, mL (baseline) | 1391.5 ± 66.0 | 1363.3 ± 73.3 | 0.125 |
| Use of CVDs drugs (anytime) | 8 (31%) | 26 (62%) | 0.024 |
| Use of CNS drugs (anytime) | 5 (19%) | 15 (36%) | 0.179 |
| Atrophy rate | 0.43 ± 0.25 | 1.46 ± 0.62 | <0.0001 |

**Table 2.** Biomarkers selected for ROC-based modelling of progression in MCI. Top variables were selected from platform-specific OPLS-DA models and evaluated individually using univariate ROC analysis to assess their ability to discriminate between stable and progressing MCI. Shown are area under the curve (AUC) values with 95% confidence intervals (CI) and t-test *p*-values. Features include metabolites from anion exchange chromatography-mass spectrometry (AEC-MS), reversed-phase liquid chromatography-mass spectrometry (RPLC-MS), and nuclear magnetic resonance (NMR; AXINON, CPMG, zgpr30), along with three targeted plasma proteins (GFAP, NfL, and pTau181). All variables were subsequently included in multivariable ROC modelling to identify the optimal prognostic biomarker panel. LDLA-c (cholesterol content in large low-density lipoprotein particles), LDLC-c (cholesterol content in central low-density lipoprotein particles), LLDL-p (particle concentration of large low-density lipoprotein), LVLDL-p (particle concentration of large very-low-density lipoprotein), VLDL-s (very-low-density lipoprotein - small subclass), GlycA/B (NMR glycoprotein signals).

| Variable | Platform | AUC | AUC<br>95% CI | *p* value |
| --- | --- | --- | --- | --- |
| 2-Hydroxyhexanoic acid | AEC-MS neg. ion mode | 0.75 | 0.63-0.88 | 0.027 |
| 5-Methoxyindoleacetate | RPLC-MS pos. ion mode | 0.56 | 0.41-0.71 | 0.072 |
| Alanine | AXINON | 0.65 | 0.51-0.79 | 0.106 |
| Butyrylcarnitine | RPLC-MS pos. ion mode | 0.62 | 0.49-0.76 | 0.058 |
| Creatinine | AXINON | 0.57 | 0.43-0.71 | 0.438 |
| Dimethylamine | AXINON | 0.54 | 0.44-0.64 | 0.432 |
| GFAP | - | 0.64 | 0.52-0.77 | 0.044 |
| Glucose region (3.40-3.48 ppm) | CPMG | 0.49 | 0.35-0.62 | 0.851 |
| Glucuronic acid | AEC-MS neg. ion mode | 0.61 | 0.46-0.75 | 0.445 |
| Glutamine (2.42-2.46 ppm) | CPMG | 0.49 | 0.36-0.63 | 0.944 |
| Glutaric acid | AEC-MS neg. ion mode | 0.53 | 0.39-0.67 | 0.568 |
| Histidine (7.04-7.06 ppm) | CPMG | 0.63 | 0.49-0.77 | 0.077 |
| Homocysteine | Targeted LC-MS | 0.55 | 0.41-0.69 | 0.310 |
| Lactate (4.10-4.12 ppm) | CPMG | 0.53 | 0.39-0.68 | 0.597 |
| Lactate/(-CH<sub>2</sub>)<sub>n</sub> (1.30-1.34 ppm) | CPMG | 0.56 | 0.42-0.70 | 0.383 |
| LDLA-c | AXINON | 0.61 | 0.47-0.75 | 0.126 |
| LDLC-c | AXINON | 0.57 | 0.42-0.71 | 0.260 |
| LLDL-p | AXINON | 0.62 | 0.48-0.76 | 0.108 |
| LVLDL-p | AXINON | 0.61 | 0.47-0.74 | 0.077 |
| Myo-inositol | AXINON | 0.60 | 0.46-0.74 | 0.217 |
| Myo-inositol/threonine/lipid (3.54-3.68 ppm) | zgpr30 | 0.64 | 0.51-0.78 | 0.089 |
| GlycA/=CH-CH<sub>2</sub>-CH<sub>2</sub>- (2.02-2.04 ppm) | zgpr30 | 0.57 | 0.43-0.71 | 0.258 |
| GlycB/=CH-CH<sub>2</sub>-CH<sub>2</sub>- /Glutamate (2.06-2.10 ppm) | zgpr30 | 0.59 | 0.46-0.73 | 0.592 |
| NfL | - | 0.61 | 0.47-0.75 | 0.180 |
| *p*-Hydroxyphenylacetic acid | AEC-MS neg. ion mode | 0.67 | 0.53-0.80 | 0.151 |
| pTau181 | - | 0.66 | 0.52-0.80 | 0.057 |
| Quinolinic acid | AEC-MS neg. ion mode | 0.52 | 0.37-0.66 | 0.694 |
| VLDL-s | AXINON | 0.60 | 0.46-0.74 | 0.125 |

### Protocol approvals, patient consent

The VITACOG trial was approved by a local NHS Research Ethics Committee (COREC 04/Q1604/100). This secondary analysis of archived plasma and serum samples adheres to STROBE guidelines; full trial details and checklists are available in Smith *et al*^19^. For the OPTIMA cohort, ethical approval was obtained from the Frenchay Research Ethics Committee (REC Ref 09/H0107/9). All participants provided written informed consent in accordance with the Declaration of Helsinki.

### UK Biobank stress-test cohort

We analysed a subset of UK Biobank participants with longitudinal MRI, plasma neurodegeneration biomarkers (pTau181, NfL, and GFAP), and Nightingale NMR metabolomics. Although future clinical trajectories remain uncertain, the cohort represents an ageing, largely asymptomatic population suitable for investigating molecular correlates of brain structural change. Brain atrophy was quantified as the annualised percentage change in total brain volume between the first and second imaging visits, using whole-brain volumes normalised for baseline, consistent with the longitudinal estimation in the VITACOG study. Participants aged ≥60 years with complete biomarker and imaging data were categorised as stable (annualised atrophy ≤0.6%; *n*=148) or progressors (≥1.0%; *n* = 75). Detailed descriptions of cohort selection, data processing, and modelling are provided in eMethods 3 and SI Fig. 1.

### MRI acquisition and atrophy rate calculation

High-resolution T1-weighted MRI scans were acquired at baseline and two-year follow-up using a 1.5T Siemens Sonata system (Oxford Centre for Clinical Magnetic Resonance Research), employing a 3D FLASH sequence with 1 mm isotropic voxels (208 coronal slices; TR=12 ms; TE=5.65 ms; flip angle =19°)^19^. Scans were performed in triplicate and averaged after alignment. Whole-brain atrophy rates were computed using SIENA, which estimates annual volume loss from longitudinal image pairs^25^. Participants were classified as stable (-0.202 to 0.716%/year) or progressors (0.761 to 3.324%/year) using a predefined threshold of 0.72%/year to differentiate normal ageing from neurodegeneration^22,28^.

### Targeted and untargeted ^1^H NMR analysis

Sample preparation and metabolomics analyses followed previously described standardised protocols. For ¹H NMR, serum samples were thawed at 4 °C and mixed with phosphate buffer (pH 7.2)^29,30^. Spectra were acquired on a 600 MHz Bruker Avance III spectrometer (Numares AG, Regensburg, Germany) using standard zgpr30 pre-saturation and CPMG pulse sequences^31^ (zgpr30 preserves macromolecular and lipoprotein signals, whereas CPMG suppresses macromolecular resonances to resolve low-molecular-weight metabolites). The NMR spectra were manually phased, baseline-corrected, and referenced to the lactate-CH₃ doublet signal (δ=1.33 ppm) using Topspin 4.1.4 (Bruker, Germany). Processed spectra were imported into ACD/NMR Processor Academic Edition 12.01 (Advanced Chemistry Development, Inc.) for further analysis. Spectral regions between 0.55-4.25 ppm and 5.20-8.50 ppm were segmented into bins of 0.02 ppm width, and the resulting integrals were sum-normalised. The bin integrals were exported for statistical analysis^29,30^. In cases where adjacent bins were selected by feature selection algorithms and corresponded to partially overlapping spectral signals, the bin integrals were aggregated and treated as a single composite feature to preserve spectral coherence and improve interpretability. Targeted NMR quantification was performed using the AXINON® platform, which applies proprietary algorithms for spectral deconvolution and absolute quantification^29^.

### Semi-targeted multi-LC-MS metabolomics analysis

Metabolite extraction for reversed-phase LC-MS (RPLC-MS) and anion-exchange LC-MS (AEC-MS) was performed on previously NMR-processed serum samples using established protocols^32^. For RPLC-MS, samples were mixed with acetonitrile (1:2.33, v/v), vortexed, centrifuged, and supernatants transferred to autosampler vials. For AEC-MS, extraction used BuOH/MeOH (1:4, v/v), followed by filtration through 10 kDa molecular weight cut-off columns. A pooled QC sample was injected throughout each sequence to monitor retention time stability, and a reference metabolite mix was analysed at the start and end of each batch to assess system performance^18^. RPLC-MS was conducted on a Waters Acquity UPLC coupled to a Xevo G2-XS QTOF mass spectrometer (Waters, UK) using gradient elution on a C18 column^32^. AEC-MS was performed using a Dionex ICS-5000+ system with an AS11-HC column and a Q Exactive Orbitrap (Thermo Fisher), with HESI II ionisation, a trap column, and a continuously regenerated electrolytic ion suppressor^33,34^. Raw data were processed in Progenesis QI (Waters, UK) for retention time alignment, peak detection, adduct deconvolution, and isotope clustering. Metabolite identification was based on accurate mass, retention time, and MS/MS spectra matched to an in-house library of authentic metabolite standards (∼150 RPLC, >450 AEC). Only compounds meeting Metabolomics Standards Initiative (MSI)^35^ Level 1 criteria and showing <30% coefficient of variation in pooled QCs were retained for downstream analysis.

### Plasma pTau181, GFAP, NfL, tHcy measurements

Plasma tHcy was measured by fluorescence polarization immunoassay using the Abbott IMx fully automated immunoassay analyser (Abbott Laboratories, Abbott Park, IL, USA). Plasma pTau181, GFAP, and NfL were measured using Single Molecule Array (Simoa) commercial Quanterix immunoassays on the HD-X platform (Quanterix, Billerica, MA, USA)^36^.

### Statistical analysis

All analyses were performed in R (R Foundation for Statistical Computing, Vienna, Austria) using custom scripts and the *ropls*, *pROC*, *caret*, and *tidyverse* packages^37^. For each analytical platform, orthogonal partial least squares discriminant analysis (OPLS-DA) was conducted on *z*-score-standardised data to model progression status in MCI, with 10-fold cross-validation repeated 100 times. Model performance was assessed on held-out test sets, and discriminative features were ranked by their average Variable Importance in Projection (VIP) scores. The top features from each platform, defined by the inflection point of the VIP score distribution (*n*=24), were selected for integrative modelling. To evaluate the prognostic value of multimodal biomarker panels, these 24 metabolites were combined with three targeted plasma proteins (GFAP, pTau181, and NfL) and tHcy, and all possible combinations of 3 to 7 features were systematically assessed using cross-validated OPLS-DA. For each combination, classification performance was quantified using both area under the receiver operating characteristic curve (ROC AUC) and mean cross-validated accuracy. Thresholds were optimised using the Youden index, and the number of orthogonal components was dynamically tuned to minimise overfitting^38^. To evaluate whether model performance exceeded chance levels, results were compared to distributions derived from 1,000 random label permutations using a two-sided Kolmogorov-Smirnov test (*p* < 0.05). The top-performing combinations, ranked by AUC and accuracy, were assembled into candidate multimodal panels for multivariable logistic regression. Final models were ranked by AUC and accuracy to identify the most prognostic and generalisable panels^39^.

Univariate analyses were conducted in R version 4.2.1. Group differences in continuous variables were assessed using unpaired two-sample t-tests, while categorical variables were compared using the Chi-square test. Statistical significance was defined as *p* < 0.05, and false discovery rate (FDR) correction using the Benjamini-Hochberg method was applied where appropriate. Data visualisation was carried out using R and complemented with figures generated in GraphPad Prism v10.

## Results

### Participant characteristics

The primary analyses were conducted in the VITACOG trial cohort, where all participants were diagnosed with MCI at baseline. To distinguish individuals showing early neurodegenerative progression from those with age-related, non-pathological changes, participants were stratified as “progressors” or “stable” MCI based on annualised whole-brain atrophy rate of 0.72% over two years as per previous reports^22,28^. Forty-two individuals were classified as progressors (atrophy rate range: 0.761-3.324%); twenty-six individuals as stable (atrophy rate range: -0.202 to 0.716%) (*p* = 3.6 E-11).

The two groups were matched at baseline across demographic and clinical characteristics, with no significant differences in age, sex, years of education, body mass index, systolic or diastolic blood pressure, initial brain volume (*p* = 0.125), or cognitive test scores (Table 1). Specifically, baseline scores did not differ for the MMSE (*p* = 0.450) or the Telephone Interview for Cognitive Status-Modified (TICS-M; *p* = 0.491) scores (Table 1). A greater proportion of progressors had received therapy for cardiovascular disease (62% vs. 31%, *p* = 0.024). However, comparisons across subgroups indicated that cardiovascular treatment was not associated with significant differences in baseline biomarkers or model performance (SI Fig. 2-5).

### Community-based analyses reveal weak coupling of protein biomarkers to brain atrophy

To complement the VITACOG analyses, we examined a community-based ageing cohort from the UK Biobank with longitudinal MRI enabling calculation of annualised cerebral atrophy rates, alongside plasma neurodegeneration markers (pTau181, GFAP, and NfL) and Nightingale NMR metabolomics (SI Fig. 1). Only six participants had a recorded diagnosis of cognitive impairment or dementia, indicating a predominantly asymptomatic ageing population. Protein biomarker distributions differed markedly between the population-based UK Biobank cohort and the clinically defined VITACOG trial. Among UK Biobank participants aged ≥60 years (*n*=311), mean concentrations were 2.3 pg/mL for pTau181, 14.0 pg/mL for NfL, and 93.0 pg/mL for GFAP (SI Tables 3-5). In VITACOG participants with MCI, mean concentrations were significantly higher - 16.3, 22.3, and 136.1 pg/mL for pTau181, NfL, and GFAP, respectively (*p* < 0.0001). The *p*-values were identical when restricting the UKB analysis to participants aged ≥70 years, but the ≥60 group was used to ensure reliable sample size (SI Tables 3-5; Fig. 1A, B).

**Figure 1.**
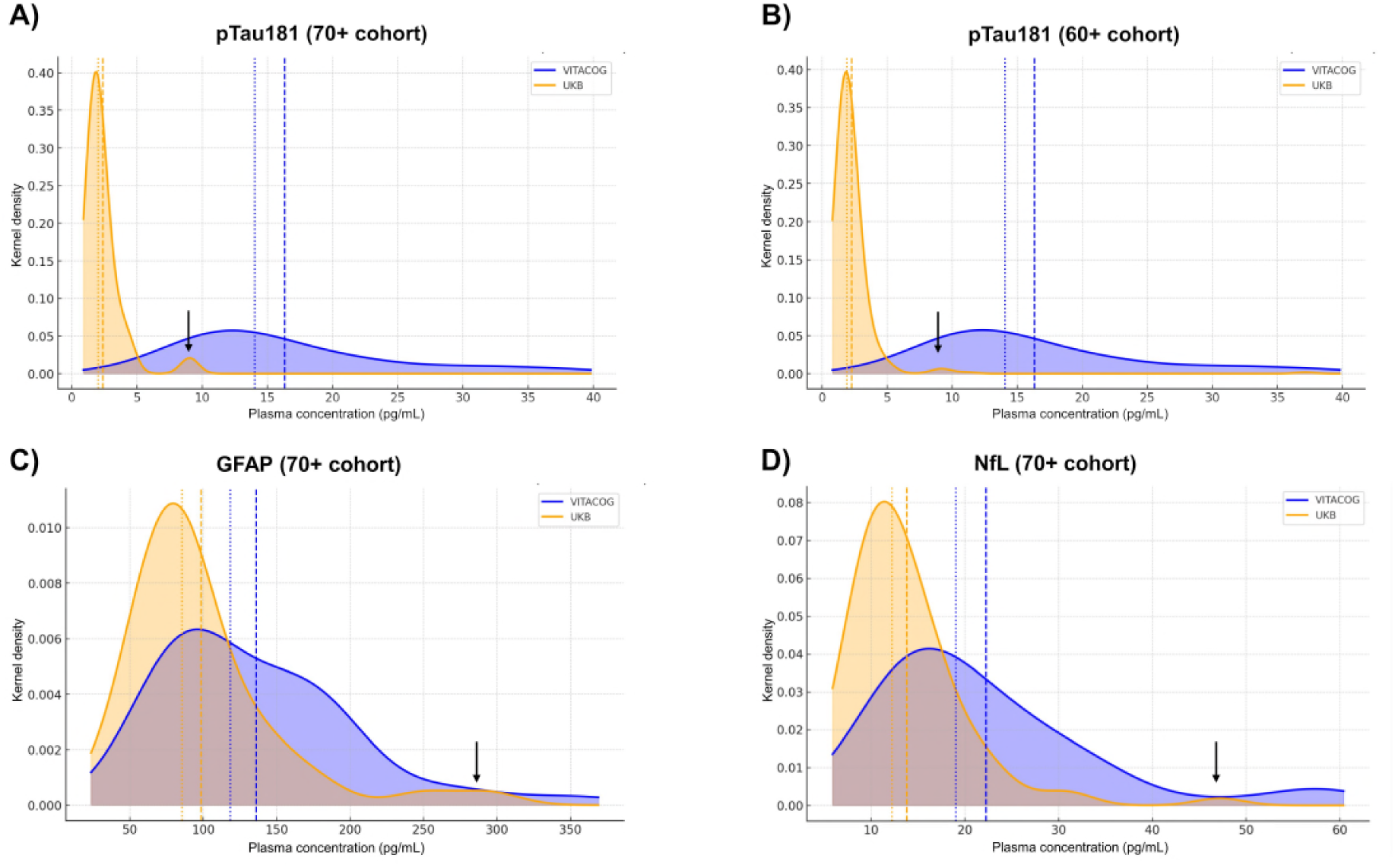
Community-based distributions of plasma neurodegeneration biomarkers reveal limited coupling to structural brain change. Kernel density plots show plasma concentrations of pTau181 **(A–B)**, GFAP **(C)**, and NfL **(D)** in VITACOG (blue) and UK Biobank (orange). Dashed vertical lines indicate mean values, and dotted lines indicate medians for each cohort. UK Biobank distributions were similar between participants aged ≥60 and ≥70 years, whereas VITACOG participants with MCI exhibited consistently higher values. Note that a subset of UK Biobank participants (5.5%; indicated by black arrows) showed markedly elevated plasma concentrations of pTau181, GFAP, or NfL (one or more biomarkers); however, none of these individuals had evidence of AD or other CNS pathology during longitudinal follow-up.

Density plots revealed distinct biomarker distributions between UK Biobank and VITACOG (Fig. 1). In UK Biobank, distributions for participants aged ≥60 and ≥70 years were nearly identical, indicating stability across late-life strata. By contrast, VITACOG participants showed right-shifted distributions with higher concentrations of all three markers, most prominently pTau181 (Fig. 1A, B). GFAP (Fig. 1C) and NfL (Fig. 1D) were also significantly higher in VITACOG (*p* < 0.0001) but exhibited greater overlap, likely reflecting modulation by diverse CNS and systemic processes, whereas pTau181 elevations appeared more specific to neurodegenerative pathology and were strongly associated with MCI in VITACOG. These patterns highlight both the heterogeneity of biomarker expression in population cohorts and the relative specificity of pTau181 for incipient neurodegeneration.

A small subset of UK Biobank participants (5.5% across all three markers) exhibited elevated concentrations of one or more plasma biomarkers of neurodegeneration (Fig. 1). Examination of ICD-10 codes revealed comorbidities such as cardiovascular, renal, pulmonary disease, or cancer, but no neurological diagnoses were reported. Outlier profiles were characterised by elevated GFAP or NfL, with or without concurrent increases in pTau181. These heterogeneous patterns likely reflect systemic rather than neurodegenerative processes, highlighting the challenge of identifying prodromal AD within community-based cohorts.

### Plasma pTau181, GFAP, and NfL show limited value for prognostication of brain atrophy in MCI

In the VITACOG MCI cohort we evaluated the performance of pTau181, GFAP, and NfL in discriminating progressors from stable individuals. ROC analysis (Fig. 2A) indicated that pTau181 exhibited the highest discriminative performance (AUC = 0.66), followed by GFAP (0.64) and NfL (0.61). Although these values reflect only modest discriminatory power in isolation, they provide a benchmark for subsequent multimodal analyses integrating metabolomic data.

**Figure 2.**
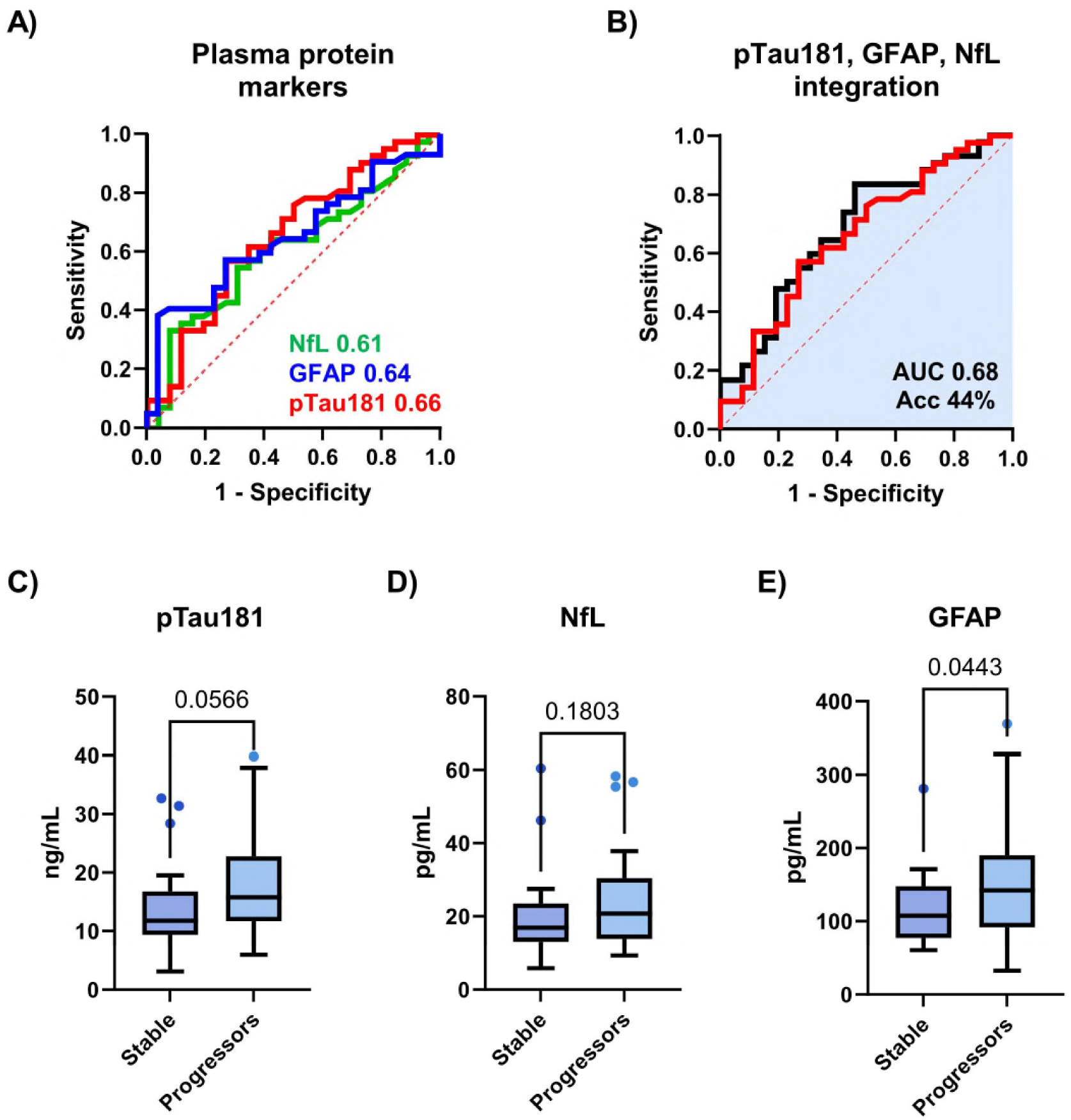
Plasma protein biomarkers show limited discrimination between stable and progressive MCI. **(A)** ROC curve analysis for individual biomarkers demonstrates modest discriminatory power, with pTau181 achieving the highest AUC (0.659), followed by GFAP (0.640) and NfL (0.610). **(B)** A multivariate model combining all three markers offers only marginal improvement (AUC 0.679; accuracy 44.3%). **(C-E)** Boxplots illustrate group-level differences, with GFAP significantly elevated in progressors (*p* = 0.0443), pTau181 showing a near-significant trend (*p* = 0.0566), and no difference observed for NfL (*p* = 0.1803).

Combining all three proteins in a logistic regression model produced a modest improvement (AUC = 0.68; Fig. 2B) with an overall classification accuracy of 44.3%. Performance remained particularly limited for clinically stable individuals, with correct classification in 19.2% for pTau181, 23.1% for GFAP, and 42.3% for the combined model. Univariate analyses confirmed group-level differences consistent with their biological roles (Fig. 2C-E). Plasma GFAP was significantly elevated in progressors (*p* = 0.0443), consistent with astroglial activation in early disease. pTau181 showed a near-significant trend towards higher levels (*p* = 0.0566), whereas NfL did not differ significantly between groups (*p* = 0.1803). Taken together, these findings reinforce that while GFAP and pTau181 capture elements of early neurodegenerative change, their effect sizes are modest. This rationale guided the evaluation of integrated protein-metabolite models to enhance prognostic accuracy.

### Integration of plasma proteins with NMR metabolomics enhances prognostication in MCI

Given the limited prognostic performance of plasma proteins alone, we next explored whether integrating complementary metabolic information could improve prognostication of brain atrophy in MCI.

To ensure balanced feature selection across modalities, OPLS-DA was performed separately for each platform (NMR CPMG, zgpr30, AXINON, and LC-MS restricted to 118 identified metabolites). Top-ranking variables by VIP score (Table 2) were then combined with pTau181, GFAP, and NfL in multivariable logistic regression models, enabling systematic evaluation of whether cross-platform integration enhances the specificity and accuracy of progression models beyond protein-based approaches. Integrated modelling revealed that compact three-variable panels combining one protein (typically GFAP or pTau181) with two NMR-derived features already outperformed protein-only models (AUC = 0.73; Fig. 3A, B). The most frequent contributors included histidine (7.04-7.06 ppm), glycoprotein acetyl groups (GlycA; 2.02-2.04 ppm), and overlapping lipid, threonine, and myo-inositol resonances (3.54-3.68 ppm). The highest accuracies (∼66%) were observed for models integrating histidine with both pTau181 and GFAP or NfL, underscoring the complementary prognostic value of protein-metabolite integration.

**Figure 3.**
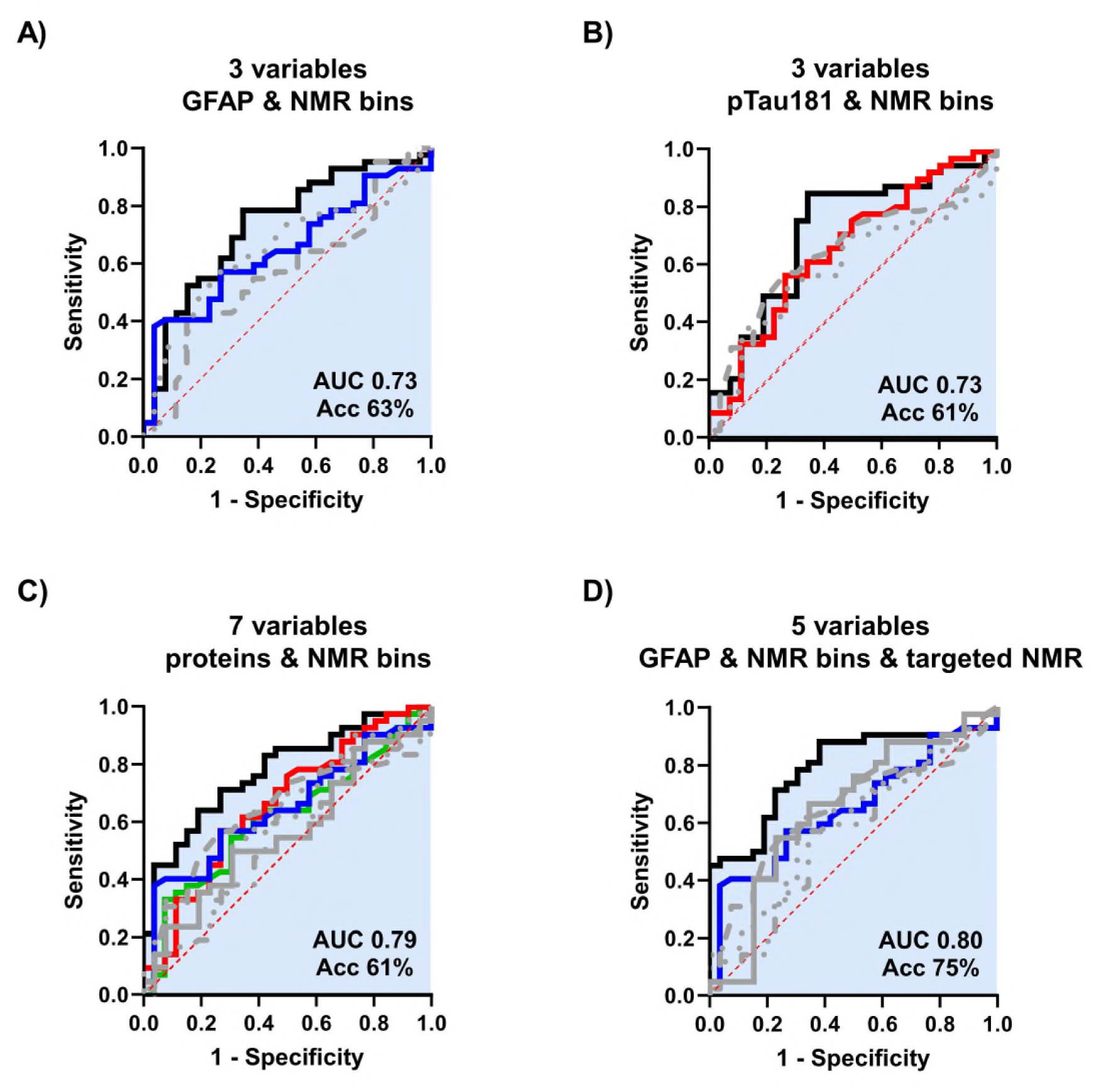
Integration of plasma proteins with NMR-derived features improves classification of MCI progression. **(A)** Model combining GFAP (blue solid line), a broad zgpr30 bin (3.54-3.68 ppm, dotted line), and GlycA (2.02-2.04 ppm, dashed line); AUC 0.73, accuracy 63%. **(B)** Model combining pTau181 (red solid line), GlycB/Glutamate (2.06-2.10 ppm, dotted line), and the zgpr30 bin (3.54-3.68 ppm, dashed line); AUC 0.73, accuracy 61%. **(C)** Model using GFAP (blue solid line), pTau181 (red solid line), NfL (green solid line), lactate (4.10-4.12 ppm, dash-dot line), histidine (7.04-7.06 ppm, dotted line), zgpr30 bin (3.54-3.68 ppm, dashed line), and lactate/alkyl chains (1.30-1.34 ppm, long dash line); AUC 0.79, accuracy 61%. **(D)** Model incorporating GFAP (blue solid line), LDLC-c (dash-dot line), LVLDL-p (dotted line), zgpr30 bin (3.54-3.68 ppm, dashed line), and alanine (long dash line); AUC 0.80, accuracy 75%.

Notably, increasing model complexity yielded only marginal performance gains. The four-variable model achieved an AUC of 0.77 with an accuracy of only 63%, while the five-, six-, and seven-variable models resulted in modest increases in AUC (up to 0.79) but plateaued in accuracy (60-62%; Fig. 3C). These results suggest that while additional features can marginally enhance sensitivity (AUC), they do not necessarily improve overall classification accuracy, underscoring the importance of model parsimony in achieving translational utility. Features consistently retained across high-performing models included plasma proteins (particularly GFAP and pTau181), the histidine resonance, and zgpr30 bins encompassing lipoprotein- and myo-inositol-related regions. Multivariable modelling further revealed additive effects of NMR features that were not apparent in univariate analyses (Table 2).

Incorporation of AXINON®-derived lipid measures further improved model interpretability and performance. A three-variable model comprising GFAP, the zgpr30 bin at 3.54-3.68 ppm, and large VLDL particle concentration (LVLDL-p) achieved an AUC of 0.80 with 73% accuracy. Replacing GFAP with pTau181 yielded comparable performance (AUC = 0.77, accuracy 72%), while adding both proteins in a four-variable model conferred no further gain. The optimal five-variable model, including GFAP, alanine, LDL cholesterol content (LDLC-c), LVLDL-p, and the zgpr30 bin, achieved an AUC of 0.80 and the highest observed accuracy (75%; Fig. 3D). Expanding to six- or seven-variable models modestly increased AUC (0.83-0.84) but reduced or plateaued accuracy, reinforcing the value of concise panels.

To assess model robustness in a broader, population-based context, we applied the same analytical framework to the UK Biobank cohort as a population-level stress test. Participants were classified as stable (*n*=148; ≤0.6% annual atrophy) or progressors (*n*=75; ≥1.0%) based on MRI-derived cerebral atrophy rates (SI Fig. 1). Cross-validated models trained on serum metabolomics and plasma neurodegeneration markers achieved moderate discrimination. Metabolite-only models reached a maximum accuracy of 66% (AUC = 0.57), driven largely by lipid-related features (VLDL cholesterol, triglycerides, HDL cholesterol, HDL particle diameter, and lactate). A five-variable model optimised for discrimination (VLDL cholesterol, tyrosine, triglycerides, LDL particle diameter, and NfL) achieved an AUC of 0.68 (59% accuracy) (SI Fig. 6A). The inclusion of protein biomarkers improved performance, with a seven-variable model combining NfL and six metabolites (VLDL cholesterol, triglycerides, valine, tyrosine, glutamine, and LDL particle diameter) yielding the highest AUC (0.70; accuracy = 58%) (SI Fig. 6B). Comparable accuracy was obtained with GFAP (AUC = 0.68) and pTau181 (AUC = 0.67), and the highest accuracy (64%) was observed for a combined pTau181-GFAP model (AUC = 0.60) (SI Fig. 6C). Although biomarker signals were diffuse in this ageing, largely non-MCI cohort, the integration of neurodegeneration markers with selected metabolites modestly enhanced discrimination, demonstrating the resilience of the multimodal framework in a population-based setting.

### AUC 0.91 achieved through LC-MS integration and broader metabolite coverage

Together, AEC-MS and RPLC-MS substantially expanded the range of metabolites detectable in blood samples. To determine whether this broader coverage could further improve predictive accuracy, LC-MS datasets were incorporated into the analysis. A three-variable model comprising 2-hydroxyhexanoic acid (AEC-MS), LVLDL-p, and the zgpr30 NMR bin achieved an AUC of 0.80 with 68% accuracy. Performance was improved when protein markers were added: a panel including pTau181, butyrylcarnitine (RPLC-MS), and 2-hydroxyhexanoic acid yielded an AUC of 0.79 and 75% accuracy, which increased to 0.85 and 77% after addition of histidine. Incorporation of glucuronic acid further increased the AUC to 0.88 while maintaining accuracy, whereas substitution with AXINON-derived myo-inositol produced an AUC of 0.87 and the highest accuracy (80%; Fig. 4A).

**Figure 4.**
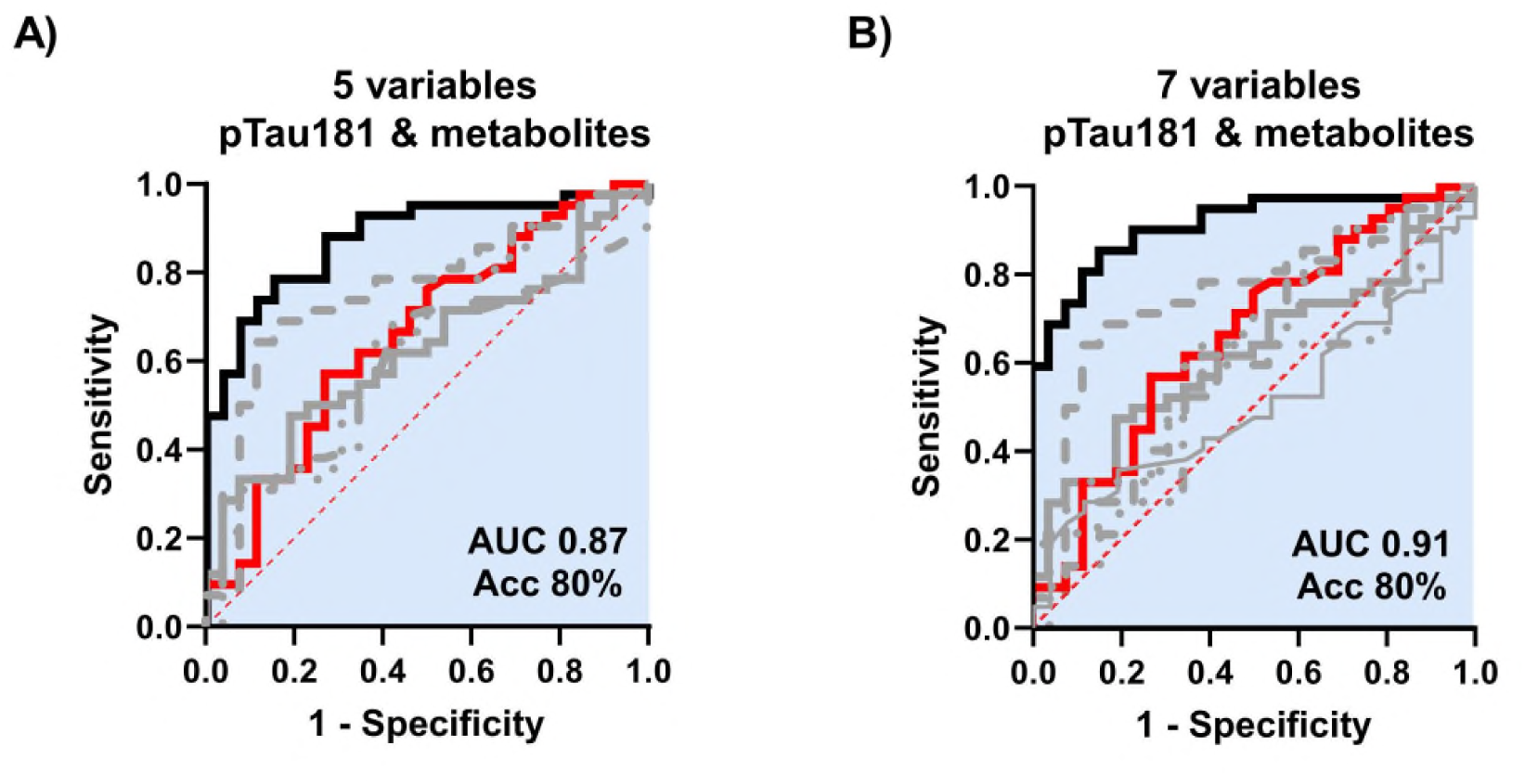
Multi-omic ROC models integrating plasma pTau181 with NMR and LC-MS metabolomic features improve discrimination of progressive MCI. **(A)** A five-variable model combining pTau181 (red solid line) with myo-inositol (AXINON, dash-dot line), 2-hydroxyhexanoic acid (AEC-MS, dashed line), butyrylcarnitine (RPLC-MS, long dash line), and histidine (CPMG, dotted line) achieved an AUC of 0.87 and accuracy of 80%. **(B)** A seven-variable model combining pTau181 (red solid line) with glucuronic acid (AEC-MS, dash-dot line), 2-hydroxyhexanoic acid (AEC-MS, dashed line), butyrylcarnitine (RPLC-MS, bold long dash line), histidine (CPMG, dotted line), glutaric acid (AEC-MS, triple-dot-dash line), and glutamine (CPMG, thin long dash line) reached an AUC of 0.91 and accuracy of 80%.

The seven-variable multi-omics models achieved the highest overall performance. A panel combining pTau181 with butyrylcarnitine, 2-hydroxyhexanoic acid, glucuronic acid, glutaric acid, histidine, and glutamate reached an AUC of 0.91 with 80% accuracy (Fig. 4B). A complementary model in which histidine and glutamate were replaced by creatinine and the zgpr30 3.54-3.68 ppm region achieved the highest accuracy (83%) with an AUC of 0.88. These extended panels improved classification of clinically stable individuals (73% correctly classified) and addressed a key limitation of protein-only models.

Independent validation of the metabolomics (NMR and LC-MS) component in the OPTIMA cohort confirmed the biological relevance and reproducibility of the VITACOG-derived signatures. When reconstructed using OPTIMA serum data, the two six-metabolite alone models achieved AUCs of 0.81 and 0.80 in *post-mortem*-confirmed AD cases (*n* = 36), closely matching the 0.85-0.86 observed in the six-metabolite alone VITACOG model (SI Fig. 7). In a broader OPTIMA subset encompassing mixed dementia cases (AD and VaD; *n* = 61), the same panels yielded AUCs of 0.75 and 0.72 (SI Fig. 8). Prognostic accuracy for MMSE decline was slightly higher in confirmed AD than in mixed-pathology cases but remained robust across both groups.

We next assessed plasma tHcy, an established biomarker of dementia risk. Although tHcy alone showed limited discriminatory performance (AUC of 0.55 in the VITACOG cohort; SI Fig. 9A), its inclusion modestly enhanced the performance of integrated models: accuracy increased from 44% to 58% in a four-variable panel combining proteins and metabolites, and to 78% in a five-variable panel, matching the best untargeted metabolomics models (SI Figs. 9B-D). These results indicate that even a single targeted biochemical measure can enhance classification when integrated with orthogonal data.

### Differential associations between circulating biomarkers and longitudinal brain atrophy in stable versus progressive MCI

To explore links between circulating markers and structural change, we evaluated correlations with longitudinal brain atrophy in stable versus progressive MCI in the VITACOG cohort using within-group Pearson correlations (Fig. 5A-D). In both groups, pTau181 showed modest positive associations with atrophy, consistent with its established role as a marker of tau pathology. In progressors, the strength of correlations varied markedly across biomarker classes. tHcy emerged as the strongest correlate of atrophy (Fig. 5B), surpassing pTau181 and all individual metabolites, despite its limited discriminative performance in univariate ROC analyses, where the AUC remained considerably lower than for pTau181 or the multi-analyte panels. When focusing on the seven-variable model (pTau181, butyrylcarnitine, 2-hydroxyhexanoic acid, glucuronic acid, glutaric acid, glutamine, and histidine), distinct correlation patterns emerged. In progressors (Fig. 5D), atrophy clustered most strongly with energy metabolism-related metabolites (butyrylcarnitine, glutaric acid, 2-hydroxyhexanoic acid), whereas in stable cases (Fig. 5C) these associations were weaker and more diffuse. Glucuronic acid and histidine showed weaker correlations with atrophy across both groups, suggesting their contributions reflect interaction effects captured only in multivariate models rather than direct linear associations.

**Figure 5.**
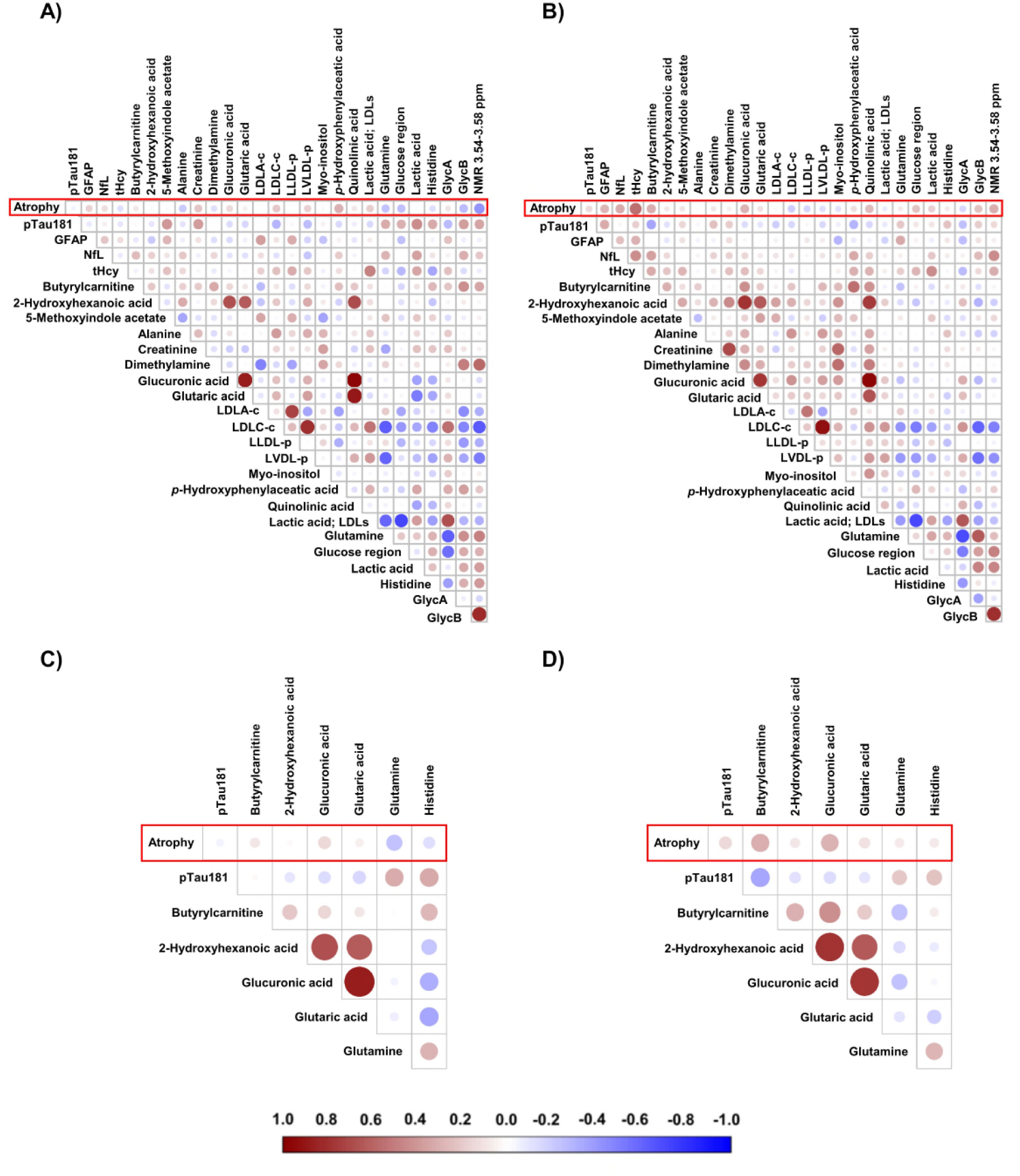
Multi-omic correlation structure linking plasma biomarkers, metabolites, and brain atrophy in stable and progressive MCI. Correlation matrices illustrate associations between atrophy rate, plasma protein biomarkers (pTau181, GFAP, NfL, tHcy), and selected metabolite and lipoprotein features. Panels **A** and **C** depict individuals with stable MCI, while panels **B** and **D** represent MCI progressors. **(A–B)** Global correlation heatmaps across all measured variables, with colour intensity and circle size proportional to Pearson’s correlation coefficients (red = positive, blue = negative). **(C–D)** Focused correlation panels for the seven-variable model comprising pTau181, butyrylcarnitine, 2-hydroxyhexanoic acid, glucuronic acid, glutaric acid, glutamine, and histidine, alongside brain atrophy (rate). In progressors, tHcy exhibited a striking positive correlation with atrophy, stronger than that observed for pTau181 or any individual metabolite, despite its low discriminative power in univariate ROC analysis. *Note:* These findings highlight that while classical biomarkers such as pTau181 contribute to prognostication, certain metabolic and vascular-related measures, including tHcy, show unexpectedly strong associations with structural decline, underscoring the value of multi-omics integration for disease progression modelling.

Together, these findings show that integration of plasma protein biomarkers with NMR- and LC-MS-based metabolomic features consistently improved predictive performance compared with proteins or metabolites alone. Compact multimodal panels combining pTau181 with metabolites related to mitochondrial, detoxification, and amino acid pathways achieved the highest accuracy (up to 80%) and AUC values of 0.91. Larger models occasionally increased AUC but did not enhance accuracy, highlighting the value of parsimonious designs. These results demonstrate that integration of orthogonal biochemical modalities enhances the robustness and generalisability of prognostic models for MCI progression.

## Discussion

Multimodal integration of plasma proteins and metabolomic profiles represents a significant advance toward improving the prognostication of MCI progression. Compact multi-omic panels combining pTau181, GFAP, and NfL with selected metabolites achieved ∼80% accuracy and AUCs >0.9, outperforming single-marker models. These results highlight the value of a multi-omics framework to capture the complex, mixed-pathology nature of early neurodegeneration and to complement, rather than replace, emerging plasma tau assays.

Plasma pTau181 levels increase progressively with cognitive decline and correlate with CSF pTau181, amyloid-β, and tau PET imaging^14^. Yet the prognostic power of pTau181, total tau^40^, or pTau217^8,9^ is variable, as these assays primarily reflect tau pathology and thus fail to capture the overall disease burden^14^. Similarly, GFAP and NfL have emerged as promising but non-specific blood biomarkers of neurodegeneration, reflecting astroglial activation and axonal injury^41,42^. Their concentrations are also age-dependent, and their release into circulation follows a non-linear pattern after neuronal injury^43^.The present findings indicate that while pTau181, GFAP, and NfL individually offer only modest discrimination, their integration with NMR- and LC-MS-derived metabolomic features substantially enhances prognostic classification of stable versus progressive MCI. Building on a well-characterised cohort, this study benefits from a robust design combining three complementary metabolomics platforms with state-of-the-art protein biomarkers, leveraging randomised controlled trial participants for model training and validating findings across both neuropathologically confirmed and population-based cohorts.

Interestingly, the prognostic performance of the multi-omics model in the UK Biobank was lower than in the discovery cohorts, but these analyses provide important context. Unlike VITACOG, which enrolled individuals with MCI at high risk of progression^19^, UK Biobank represents a heterogeneous ageing population in which only a minority are expected to present with prodromal dementia. The weaker biomarker-atrophy associations observed in this setting therefore emphasise that MCI is a biologically distinct state, characterised by stronger coupling between plasma markers and structural brain change. Accordingly, the UK Biobank analyses act as a population “stress-test,” reinforcing our findings that biomarker elevations are common in community cohorts but gain specificity in carefully phenotyped, at-risk groups. Underscoring this, elevated plasma pTau181, GFAP, and NfL can occur in individuals without evidence of CNS disease, even after longitudinal follow-up. This highlights the challenge of high sensitivity but limited specificity when such markers are applied indiscriminately at the population level. For healthcare systems, this raises a tangible risk of false positives, unnecessary anxiety, and overburdening of memory services. By contrast, our findings demonstrate that in the right setting, multi-omic panels restore specificity, capturing prodromal neurodegenerative processes underlying cognitive decline while filtering out non-specific systemic signals.

We next examined the neuropathologically confirmed OPTIMA cohort^26^ to validate whether the metabolic features identified in VITACOG reflected core processes of neurodegeneration. MCI is a heterogeneous clinical state rather than a uniform precursor of AD; pure AD pathology is rare, and most cases involve overlapping amyloid, tau, vascular, metabolic, and inflammatory mechanisms^26^. This pathological diversity limits the prognostic value of single protein biomarkers, which largely capture tau or glial injury. Metabolomic profiling, by contrast, detects a wider range of biochemical and systemic disturbances, providing orthogonal insight into disease mechanisms. In the OPTIMA cohort, prognostic accuracy for cognitive decline (MMSE change) was slightly higher in the subset with neuropathologically confirmed AD compared with the group exhibiting mixed pathology, although performance remained robust across both. This pattern suggests that the identified metabolites are particularly sensitive to AD-related neurodegeneration, yet retain discriminatory power in the presence of additional vascular or metabolic contributions. The ability of these signatures to generalise across individuals with coexisting pathologies reinforces their value as markers of overall neurodegenerative burden rather than disease-specific pathology, reflecting the biological complexity that characterises MCI and late-life cognitive decline.

Consistent with prior metabolomic studies^18,44^, our analyses reveal reproducible disturbances in amino acid metabolism (particularly glutamate, glutamine, and histidine) implicating excitotoxicity, impaired neurotransmission, and a dysregulated glutamate-glutamine cycle^45,46^, recognised drivers of synaptic dysfunction^47^. Alterations in lipid transport and lipoprotein subclasses, especially HDL and VLDL, were also evident, while the identification of hydroxy- and dicarboxylic acids supports growing evidence that defects in mitochondrial energy metabolism, including lactate and α-ketoglutarate^45^, contribute to AD^48^ pathogenesis. The strongest prognostic performance in our study was achieved with a seven-variable model integrating pTau181 with glucuronic acid, 2-hydroxyhexanoic acid, butyrylcarnitine, histidine, glutaric acid, and glutamine (AUC 0.91; accuracy 80%). Collectively, these metabolites converge on biologically plausible pathways encompassing neurotransmission, energy metabolism, redox balance, and detoxification, reflecting both central and systemic components of disease. Glutamine and histidine highlight disturbances in amino acid turnover, consistent with impaired glutamate-glutamine cycling, excitotoxicity, and reduced histaminergic signalling^47^. Glutaric acid and 2-hydroxyhexanoic acid indicate mitochondrial and peroxisomal dysfunction, while butyrylcarnitine reflects impaired β-oxidation, in line with prior reports of altered acylcarnitine profiles in MCI and AD^49^. Glucuronic acid, by contrast, marks detoxification and clearance, with impaired glucuronidation in AD linked to defective removal of lipid peroxidation products and xenobiotics^50^. Together, these metabolites delineate a systemic signature that complements tau-specific markers, providing a mechanistic rationale for the superior prognostic power of multi-omics integration. Importantly, these pathways extend beyond diagnostics, as one-carbon metabolism, lipid dysregulation, and oxidative stress represent therapeutic entry points, with interventions such as tHcy-lowering by B-vitamins^19^, lipid-modulating strategies, and antioxidant approaches already showing promise^51^.

Although tHcy alone showed limited discriminatory power, its inclusion alongside proteins and metabolites consistently improved model performance. This aligns with evidence from the VITACOG trial, where elevated tHcy correlated with accelerated brain atrophy and cognitive decline^19,52^. Mechanistically, tHcy has been linked to impaired methylation capacity^53^, vascular dysfunction^54^, oxidative stress^55^, and direct neurotoxicity^18^, all plausible contributors to AD progression. Notably, incorporating tHcy into multi-omics models illustrates how targeted, mechanistically grounded biomarkers can augment untargeted metabolomics and protein measures. Another key observation is that relatively parsimonious models demonstrate superior performance. Panels with only three to five features frequently achieved ≥75% accuracy and AUCs approaching 0.9, while larger models showed diminishing returns and reduced generalisability. Such compact models are well suited for clinical translation, minimising assay complexity while retaining interpretability. The integration of multiple analytical platforms further strengthens performance, reflecting the orthogonality of their components, proteins indexing neurodegenerative pathology and metabolites capturing systemic metabolic responses.

### Limitations

Although the sample sizes within individual cohorts were modest, the inclusion of three independent datasets strengthens reproducibility and generalisability. This multi-cohort approach, integrating randomised trial participants, neuropathologically confirmed cases, and community-based samples, enhances diversity and representativeness. However, harmonisation across analytical platforms, particularly between NMR and LC-MS, introduces variability that highlights the challenges of integrating different technologies and underscores the need for further standardisation. While the study used pTau181, the framework is assay-agnostic and could be adapted for emerging tau markers such as pTau217. The absence of protein measures in the OPTIMA cohort limits cross-modality comparison. Despite these constraints, the convergence of findings across cohorts and platforms strongly supports the utility of integrated metabolomic-proteomic signatures in modelling disease progression. However, to confirm the prognostic value of the identified multi-omic panels and their stability across diverse populations, larger, prospective studies incorporating longitudinal cognitive and imaging endpoints will be essential

## Conclusion

As disease-modifying therapies for AD enter clinical use, scalable blood-based tools for early risk stratification are required. Our findings show that compact multi-omic panels integrating plasma proteins with metabolomic features capture both central and systemic components of early neurodegeneration, achieving high prognostic accuracy in MCI. By complementing rather than replacing tau assays, this framework captures overall disease burden while enhancing biological specificity. Importantly, while NMR and LC-MS are highly sensitive and informative technologies, ongoing advances in accessibility and cost-effectiveness are making them increasingly viable for broader clinical application.

## Data Availability

Anonymised data not published within this article will be made available by request from any qualified investigator.

## Contribution statement

T.K. and D.C.A. conceived and designed the study. A.D.S. and H.R. provided study oversight and coordinated patient recruitment. D.S. supervised biobank management. F.J. performed lipid assays and managed the VITACOG database. T.K. and E.P. performed the experiments, and together developed and optimised the metabolomics sample preparation protocols. T.K. processed and analysed the data. J.J.J.J.M., D.R.S., and T.K. performed UK Biobank data mining and optimised the multivariate data analysis. S.J., Q.G., and E.S. oversaw the NMR analyses. J.S.O.M. supervised the LC-MS analyses. S.M.S. oversaw MRI acquisition and performed atrophy measurements in the VITACOG cohort. A.F.J. and D.L. provided clinical input and interpretation. L.M.-G. performed the targeted protein measurements. N.J.A. and H.Z. supervised the targeted protein measurements. D.C.A. and J.S.O.M. provided overall supervision. T.K. wrote the manuscript, with critical input from D.C.A., J.S.O.M., A.F.J., D.L., J.J.J.J.M., T.O., H.R., and A.D.S. All authors reviewed and approved the final manuscript.

## Ethics approval and consent to participate

The trial, registered as VITACOG under the title *“Homocysteine and B Vitamins in Cognitive Impairment”* (ISRCTN 94410159), was conducted in accordance with the principles outlined in the Declaration of Helsinki. Ethical approval was granted by the local NHS research ethics committee (COREC 04/Q1604/100). Written informed consent was obtained from all participants prior to their enrolment in the study.

## Acknowledgements

The authors thank Prof. Kaj Blennow (Department of Psychiatry and Neurochemistry, Institute of Neuroscience and Physiology, The Sahlgrenska Academy, University of Gothenburg, Gothenburg, Sweden) for his assistance in acquiring the pTau181, GFAP, and NfL data. We also thank all MCI patients participating in the VITACOG and OPTIMA studies, and the NMR and MS Research Facility (Department of Chemistry, University of Oxford) for access to equipment and expert advice.

