## Supplementary Information for "Multi-Omics Modelling of Plasma pTau181, GFAP, and Metabolic Features Enables Risk Stratification in Prodromal Alzheimer’s Disease"

*[1]Chemistry Research Laboratory, Department of Chemistry, University of Oxford, Oxford, UK*
*[2] Kavli Institute of Nanoscience Discovery, Dorothy Crowfoot Hodgkin Building, Oxford, UK*
*[3] Physical and Theoretical Chemistry, University of Oxford, Oxford, UK*
*[4] Oxford Centre for Magnetic Resonance Research, University of Oxford, Oxford, UK*
*[5] Department of Clinical Medicine, Aarhus University, Aarhus, Denmark*
*[6] Department of Pharmacology, University of Oxford, Oxford, UK*
*[7]Oxford Centre for Integrative Neuroimaging, FMRIB, Nuffield Department of Clinical Neurosciences, University of Oxford, Oxford, UK*
*[8] Department of Pharmaceutical Biosciences, Uppsala University, Uppsala, Sweden*
*[9] numares AG, Regensburg, Germany*
*[10] Department of Psychiatry and Neurochemistry, Institute of Neuroscience and Physiology, The Sahlgrenska Academy, University of Gothenburg, Gothenburg, Sweden*
*[11]King's College London, Institute of Psychiatry, Psychology and Neuroscience, Maurice Wohl Clinical Neuroscience Institute, London, UK*
*[12]NIHR Biomedical Research Centre for Mental Health and Biomedical Research Unit for Dementia at South London and Maudsley NHS Foundation Trust, London, UK*
*[13]Centre for Age-Related Medicine, Stavanger University Hospital, Stavanger, Norway*
*[14]Department of Nutrition, Institute of Basic Medical Sciences, Faculty of Medicine University of Oslo, Oslo, Norway*
*[15] Department of Neurology, University Hospital and University of Basel, Basel, Switzerland*
*[16] Multiple Sclerosis Centre, Departments of Biomedicine and Clinical Research, University Hospital and University of Basel, Basel, Switzerland*
*[17] Research Center for Clinical Neuroimmunology and Neuroscience Basel, University Hospital and University of Basel, Switzerland*
*[18] Clinical Neurochemistry Laboratory, Sahlgrenska University Hospital, Mölndal, Sweden*
*[19] Department of Pathology and Laboratory Medicine, University of Wisconsin School of Medicine and Public Health, Madison, WI, USA*
*[20] UK Dementia Research Institute at University College London, London, UK*

****Corresponding author:***

*Tereza Kacerova*

**

*Chemistry Research Laboratory, Department of Chemistry, University of Oxford, Oxford OX1 3TA, UK*
*Kavli Institute of Nanoscience Discovery, Dorothy Crowfoot Hodgkin Building, Oxford OX1 3QU, UK*
*Physical and Theoretical Chemistry, University of Oxford, Oxford OX1 3QZ, UK*

### eMethods 1: Validation of metabolomics results in the OPTIMA cohort: *post-mortem*-confirmed Alzheimer's disease

#### Cohort description and study outline

Metabolomic findings from the discovery VITACOG cohort were validated in a subset of the Oxford Project to Investigate Memory and Ageing (OPTIMA) comprising individuals with *post-mortem*-confirmed AD. This neuropathological confirmation provides a major advantage, ensuring that the observed metabolomic patterns reflect true AD pathology. Inclusion criteria required a MMSE score ≥24 at blood collection, approximating the MCI stage in VITACOG. To mirror the longitudinal dynamics of the discovery cohort, stable participants were defined as having ΔMMSE of 0-1 and progressors as ΔMMSE of 4-13 over two years, yielding a validation cohort of 18 stable and 19 progressors (mean age ± SD: 74.7 ± 6.4 and 72.4 ± 9.6 years, respectively). No significant differences were observed in age ($p = 0.412$) or sex distribution (50% vs 58% female). The slightly lower baseline MMSE in progressors (mean difference 1.5 points; $p = 0.042$) reflects the requirement to define progression based on cognitive decline.

**SI Table 1. Demographic and clinical characteristics of OPTIMA participants (*post-mortem*-confirmed AD)** classified as stable or progressors based on the change of Mini-Mental State Examination (MMSE) scor**e.** Data are presented as mean ± standard deviation. Between-group comparisons were performed using unpaired two-tailed t-tests for continuous variables and chi-square tests for categorical variables. A *p*-value < 0.05 was considered statistically significant.

| Characteristics | Stable *n* = 18 | Progressor *n* = 19 | *p*-value |
|---|---|---|---|
| Age (baseline) | 74.72 ± 6.39 | 72.42 ± 9.59 | 0.412 |
| Sex (% female) | 9 (50.0%) | 11 (57.9%) | 0.746 |
| MMSE (baseline) | 27.67 ± 2.16 | 26.26 ± 1.74 | 0.042 |
| Delta MMSE (2 years) | 0.33 ± 0.47 | 6.79 ± 2.80 | <0.0001 |
| *Disease trajectory* | | | |
| AD | 6 | 13 | 0.098 |
| AD + CVD | 5 | 3 | |
| AD + DLB | 7 | 3 | |
| Use of CVDs drugs (anytime) | 3 | 1 | 0.340 |
| Use of CNS drugs (anytime) | 3 | 2 | 0.660 |

The MMSE is a coarse screening tool with limited dynamic range at the upper end, and baseline differences of 1-2 points fall within normal measurement error. MMSE cut-offs were chosen empirically to maximise separation between individuals with stable cognition and those showing meaningful decline, while minimising misclassification due to measurement variability. The MMSE has a known test-retest standard deviation of approximately 1.5-2.0 points in older adults, even when cognition is stable (doi:

10.1016/j.acn.2004.11.004). Accordingly, changes of 0-1 points are within normal fluctuation, whereas declines ≥4 points exceed the 95% confidence limits for test–retest variation and indicate clinically evident progression toward dementia in longitudinal MCI studies. Intermediate changes (2-3 points) were excluded to ensure analytical clarity and reduce bias from borderline cases.

This approach yielded two clearly separated groups representing stable and progressive trajectories, consistent with definitions used in previous MCI-to-AD validation studies. Importantly, the within-group standard deviation of baseline MMSE was small relative to the magnitude of change used for stratification, indicating that apparent baseline differences are trivial once longitudinal change is considered. Thus, baseline MMSE differences primarily reflect the coupling between initial status and subsequent rate of decline, rather than bias in group allocation (SI Table 1).

**eMethods 2: Validation of metabolomics results in the OPTIMA cohort: *post-mortem*-confirmed diagnosis (mixed pathology)**

### Cohort description and study outline

To further assess the generalisability of the identified metabolic signatures, validation was extended to a larger OPTIMA subset comprising individuals with mixed neuropathological diagnoses, including both AD and cases of vascular dementia (VaD). Participants met criteria for mild cognitive impairment at baseline (MMSE ≥24) and were classified as stable (*n*=35) or progressive (*n*=26) based on longitudinal cognitive trajectories over two years (ΔMMSE 0-1 vs 4-19, respectively).

No significant differences were observed in baseline age ($p$ = 0.139) or sex distribution ($p$ = 0.613) between groups, and baseline MMSE was comparable ($p$ = 0.675; SI Table 2). This subset therefore enabled evaluation of model robustness across a broader neuropathological spectrum without potential bias from baseline cognitive performance. The inclusion of mixed pathologies provides an important test of biological specificity and clinical relevance, demonstrating that the metabolic component of the multi-omic model remains informative of cognitive decline even in the presence of overlapping disease mechanisms.

**SI Table 2. Demographic and clinical characteristics of OPTIMA participants (*post-mortem* confirmed AD and vascular dementia (VaD))** classified as stable or progressors based on the change of Mini-Mental State Examination (MMSE) scor**e.** Data are presented as mean ± standard deviation. Between-group comparisons were performed using unpaired two-tailed t-tests for continuous variables and chi-square tests for categorical variables. A *p*-value < 0.05 was considered statistically significant.

| Characteristics | Stable<br>*n* = 35 | Progressor<br>*n* = 26 | *p*-value |
|---|---|---|---|
| Age (baseline) | 75.91 ± 6.62 | 73.08 ± 7.89 | 0.139 |
| Sex (% female) | 15 (42.9%) | 13 (50.0%) | 0.613 |
| MMSE (baseline) | 27.34 ± 1.79 | 27.54 ± 1.74 | 0.675 |
| Delta MMSE (2 years) | 0.34 ± 0.48 | 10.77 ± 5.35 | <0.0001 |
| *Disease trajectory* | | | |
| AD | 6 | 13 | 0.053 |
| AD + CVD | 5 | 3 | |
| AD + DLB | 7 | 3 | |
| VaD | 17 | 7 | |
| Use of CVDs drugs (anytime) | 4 | 4 | 0.713 |
| Use of CNS drugs (anytime) | 5 | 2 | 0.688 |

**eMethods 3: UK Biobank validation analyses**

To assess molecular signatures of neurodegeneration in the general population, we analysed a subset of UK Biobank participants with longitudinal MRI, plasma neurodegeneration markers, and Nightingale NMR metabolomics. Of 1,273 individuals with biomarker data, 1,114 had ≥2 MRI scans suitable for atrophy estimation, and 654 also had NMR data (Fig. S1). Only six carried a baseline diagnosis of cognitive impairment or dementia, indicating a predominantly asymptomatic ageing cohort. Brain atrophy was calculated as the annualised percentage change in total brain volume (grey and white matter; UKB field #25010) between the first and second MRI scans (second and third UKB visit). Circulating biomarkers (pTau181, NfL, GFAP) and serum metabolites were evaluated for associations with atrophy, and protein-metabolite integration was tested for added prognostic value. Because the ≥70-year subgroup (*n*=95) was too small, analyses were extended to those aged ≥60 years (*n*=311). This provided an unselected ageing population for comparison with the carefully phenotyped VITACOG MCI cohort.

Since COVID-19 infection may impact brain volume, we reviewed the ICD-10 records to identify participants with a COVID-19 diagnosis. A total of seven individuals had a COVID-19 diagnosis (U07.1 or U07.3), of whom only three were lab-confirmed cases (U07.1). Importantly, all these individuals were stable, and the presence of COVID-19 diagnoses in this subgroup represents less than 5% of the stable population. Given that these participants with confirmed COVID-19 were stable and accounted for a very small proportion of the cohort, any potential impact of COVID-19 on brain volume measurements is unlikely to have influenced the overall findings. This further supports the robustness of our results, with minimal risk of confounding.

**eMethods 3: UK Biobank validation analyses**

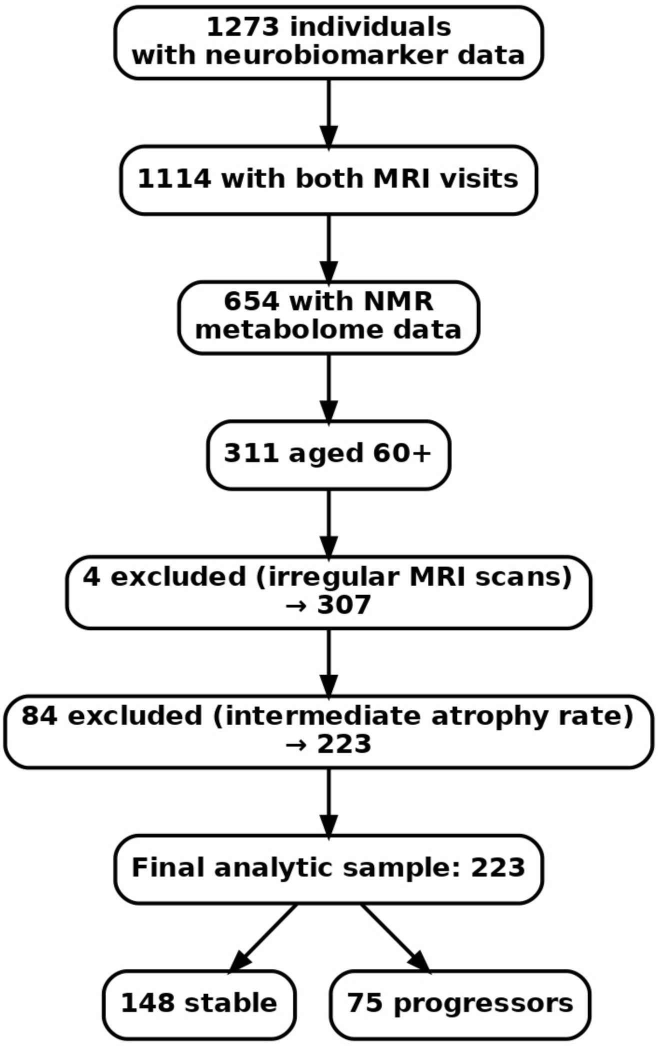


**SI Figure 1. Flow diagram of cohort selection for stress-test analysis in UK Biobank.** Of 1,273 individuals with available plasma neurobiomarkers (pTau181, NfL, GFAP), 1,114 had both MRI visits and 654 had NMR metabolomics data. Restricting to those aged ≥60 years yielded 311 participants. After excluding individuals with irregular MRI scans (*n*=4) and those with intermediate atrophy rates (*n*=84), the final analytic sample comprised 223 participants, classified as 148 stable and 75 progressors based on whole-brain atrophy rates.

Using cut-offs to separate "stable" participants (*n*=148; annualised atrophy ≤0.6%) from "progressors" (*n*=75; annualised atrophy ≥1.0%), we trained cross-validated models based on serum metabolomics alone or in combination with plasma neurodegeneration markers (pTau181, NfL, GFAP). Prognostic performance was modest: metabolite-only models achieved maximum accuracies of ~55% with AUCs up to 0.68 (Figure S2), while inclusion of protein markers marginally improved AUC (0.70) without increasing accuracy (Figure S4). An "extreme-case" analysis, contrasting the 25 individuals with the highest atrophy rates against the 25 with the lowest, yielded a classification accuracy of 54% (Figure S5).

### Impact of cardiovascular pharmacotherapy on baseline biomarkers and model performance

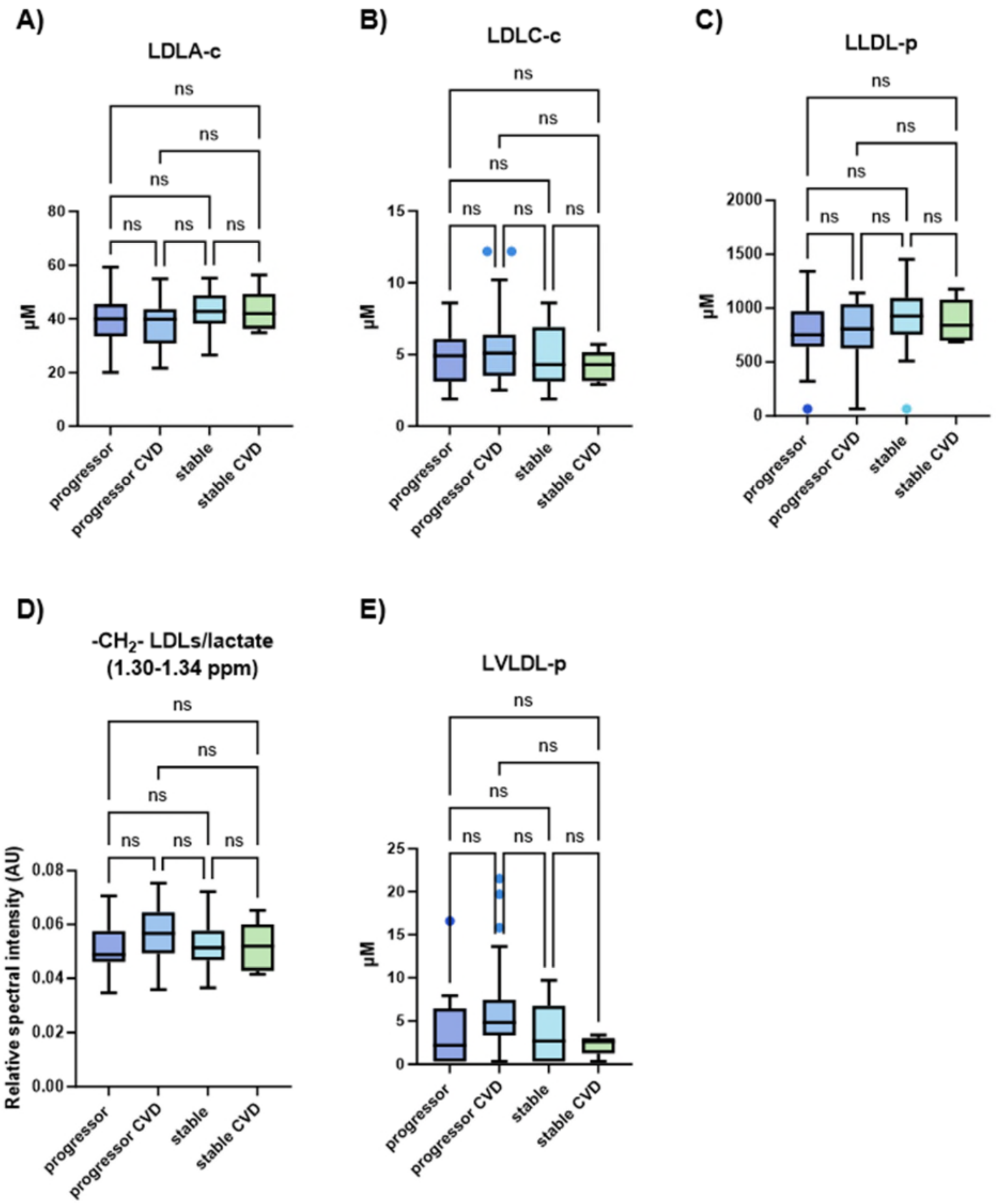


**SI Figure 2. Impact of CVD pharmacotherapy on lipoprotein and lipid-associated biomarkers.** Boxplots showing baseline levels of AXINON-derived lipoprotein parameters and NMR lipid signals across four subgroups: progressors (no CVD pharmacotherapy), progressors with CVD drugs, stable (no CVD pharmacotherapy), and stable with CVD therapy. No significant differences were observed in the biomarkers between groups (1-way ANOVA $p > 0.05$), indicating that CVD pharmacotherapy did not significantly alter baseline lipoprotein and lipid-associated signals.

To explore potential associations between cardiovascular disease (CVD) pharmacotherapy and baseline biomarker levels, we examined AXINON-derived lipoprotein parameters and NMR-derived lipid signals across the following subgroups: progressors (no CVD pharmacotherapy), progressors with CVD drugs, stable (no CVD pharmacotherapy), and stable with CVD therapy. Despite the observed marginally significant difference in CVD pharmacotherapy ($p = 0.02$) across groups, no lipoprotein signals or lipid-associated NMR bins were significantly influenced by CVD drug administration or the presence of CVD itself. One-way ANOVA revealed no significant differences ($p > 0.05$), suggesting that CVD therapy did not markedly alter the levels of these cardiovascular biomarkers at baseline (SI Fig. 2).

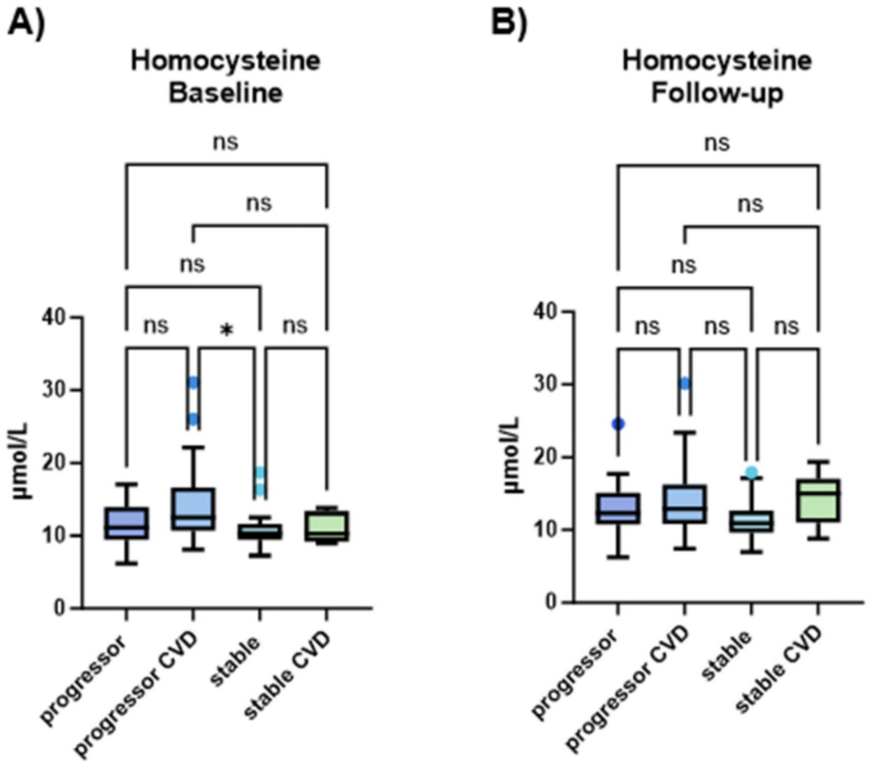


**SI Figure 3. Baseline (A) and follow-up (B) levels of total homocysteine (tHcy) across four subgroups**: progressors without CVD pharmacotherapy, progressors with CVD pharmacotherapy, stable without CVD pharmacotherapy, and stable with CVD pharmacotherapy. While no significant differences were observed between groups in response to CVD treatment, a marginally significant difference in tHcy levels was noted in two subgroups, correlating with disease progression. Statistical significance is indicated by an asterisk (*) in panel A.

While baseline and follow-up levels of tHcy, a well-established CVD biomarker, did not show significant differences in response to CVD pharmacotherapy (SI Fig. 3), a marginally significant difference was observed in two subgroups related to disease progression. This finding supports the correlation between tHcy levels and disease progression, rather than CVD treatment, and aligns with the lack of changes observed in other lipoprotein and lipid-associated biomarkers.

Finally, we examined the baseline levels of the three target proteins - pTau181, NfL, and GFAP - and found no differences related to CVD treatment (SI Fig. 4). However, GFAP exhibited statistically significant differences (*p* 0.02) associated with disease progression, as shown in Figure 2E.

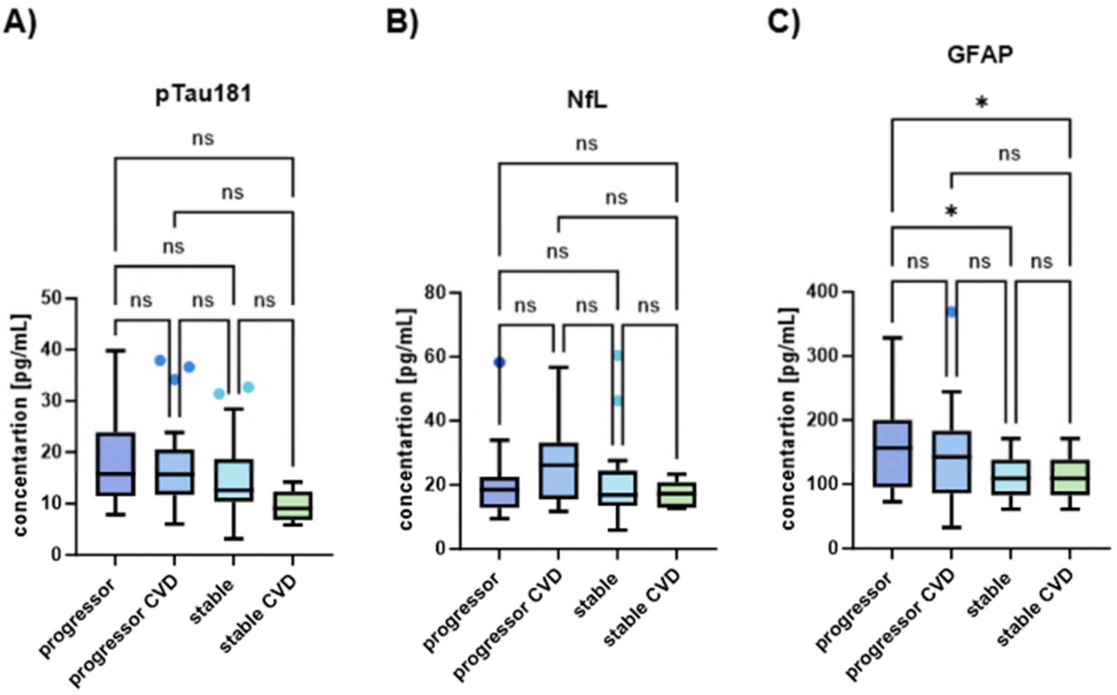


**SI Figure 4. Baseline concentrations of pTau181 (A), NfL (B), and GFAP (C) across four subgroups:** progressors without CVD pharmacotherapy, progressors with CVD pharmacotherapy, stable without CVD pharmacotherapy, and stable with CVD pharmacotherapy. No significant differences were observed in pTau181 and NfL levels across the groups. However, GFAP levels showed statistically significant differences ($p = 0.02$) in relation to disease progression (C).

Finally, we replicated the most discriminatory models to evaluate whether excluding patients taking CVD drugs would affect prognostic performance. The AUC for all models containing lipid-derived features remained comparable, with only a subtle decrease in accuracy (SI Fig. 5). This minor reduction was likely due to the reduced sample size available for model building and cross-validation rather than any significant impact of CVD treatment. These results suggest that CVD pharmacotherapy does not notably alter the prognostic performance of the model.

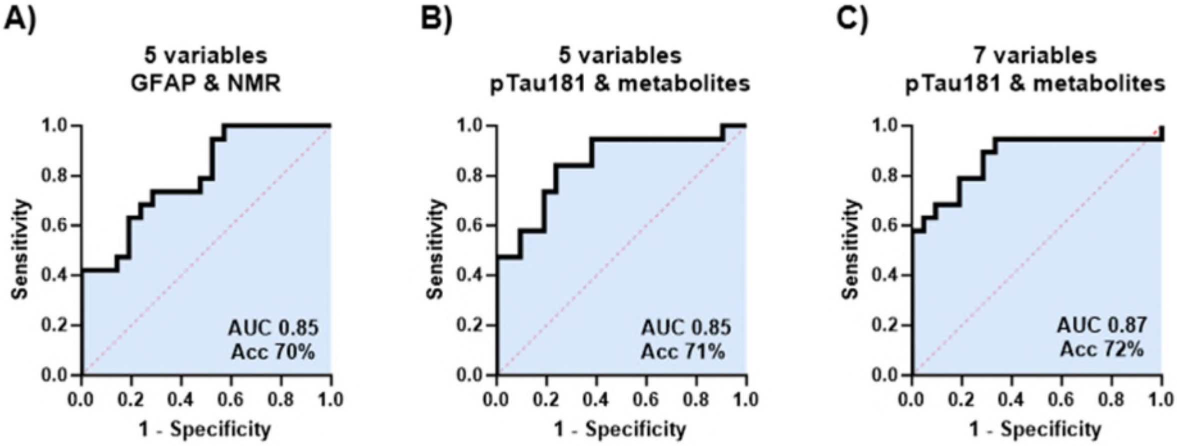


**SI Figure 5. ROC curves for the most discriminatory models, showing performance with and without inclusion of patients on CVD pharmacotherapy. (A)** displays the model using 5 variables: GFAP and four NMR-derived metabolites (refer to main text Fig. 3D); **(B)** uses 5 variables (pTau181 integrated with metabolites) - refer to main text Fig. 4A; and **(C)** uses 7 variables (pTau181 integrated with metabolites) - refer to main text Fig. 4B. Despite excluding patients on CVD treatment, the AUC remained comparable, with subtle decreases in accuracy, likely due to the reduced sample size, rather than an effect of CVD pharmacotherapy.

### Community-based analyses reveal weak coupling of biomarkers to brain atrophy

**SI Table 3. Summary statistics for plasma pTau181 in UK Biobank and VITACOG.** Descriptive statistics for pTau181 concentrations across UK Biobank participants aged ≥60 years, ≥70 years, the ≥60-year extreme atrophy subgroup, and VITACOG MCI participants.

| pTau181 | All 60+ | 60+ extremes | All 70+ | VITACOG |
|---|---|---|---|---|
| Patient count | 311 | 223 | 89 | 68 |
| Mean [pg/mL] | 2.3 | 2.3 | 2.4 | 16.3 |
| Median [pg/mL] | 1.9 | 2.0 | 2.0 | 14.1 |
| Std. dev. [pg/mL] | 2.3 | 2.6 | 1.5 | 8.3 |
| Min value [pg/mL] | 0.8 | 0.8 | 1.0 | 3.1 |
| Max value [pg/mL] | 37.1 | 37.1 | 9.4 | 39.8 |
| Range [pg/mL] | 36.3 | 36.3 | 8.4 | 36.7 |
| Lower 95% CI [pg/mL] | 2.0 | 2.0 | 2.1 | 14.3 |
| Upper 95% CI [pg/mL] | 2.5 | 2.7 | 2.7 | 18.3 |

**SI Table 4. Summary statistics for plasma NfL in UK Biobank and VITACOG.** Descriptive statistics for pTau181 concentrations across UK Biobank participants aged ≥60 years, ≥70 years, the ≥60-year extreme atrophy subgroup, and VITACOG MCI participants.

| NfL | All 60+ | 60+ extremes | All 70+ | VITACOG |
|---|---|---|---|---|
| Patient count | 311 | 223 | 89 | 68 |
| Mean [pg/mL] | 14.0 | 14.2 | 13.6 | 22.3 |
| Median [pg/mL] | 12.9 | 12.9 | 12.0 | 19.1 |
| Std. dev. [pg/mL] | 5.9 | 5.7 | 6.2 | 12.0 |
| Min value [pg/mL] | 5.1 | 5.1 | 6.0 | 5.9 |
| Max value [pg/mL] | 54.9 | 47.2 | 47.2 | 60.4 |
| Range [pg/mL] | 49.8 | 42.1 | 41.2 | 54.5 |
| Lower 95% CI [pg/mL] | 13.4 | 13.4 | 12.3 | 19.4 |
| Upper 95% CI [pg/mL] | 14.7 | 14.9 | 14.9 | 25.2 |

**SI Table 5. Summary statistics for plasma GFAP in UK Biobank and VITACOG.** Descriptive statistics for pTau181 concentrations across UK Biobank participants aged ≥60 years, ≥70 years, the ≥60-year extreme atrophy subgroup, and VITACOG MCI participants.

| GFAP | All 60+ | 60+ extremes | All 70+ | VITACOG |
| --- | --- | --- | --- | --- |
| Patient count | 311 | 223 | 89 | 68 |
| Mean [pg/mL] | 93.0 | 92.5 | 95.7 | 136.1 |
| Median [pg/mL] | 82.9 | 80.3 | 84.4 | 118.5 |
| Std. dev. [pg/mL] | 46.8 | 48.9 | 50.0 | 66.0 |
| Min value [pg/mL] | 23.6 | 31.4 | 23.6 | 32.9 |
| Max value [pg/mL] | 377.6 | 377.6 | 295.9 | 369.0 |
| Range [pg/mL] | 354.0 | 346.2 | 272.3 | 336.1 |
| Lower 95% CI [pg/mL] | 87.7 | 86.1 | 85.2 | 120.1 |
| Upper 95% CI [pg/mL] | 98.2 | 99.0 | 106.2 | 152.0 |

**Community-based analyses reveal weak coupling of biomarkers to brain atrophy**

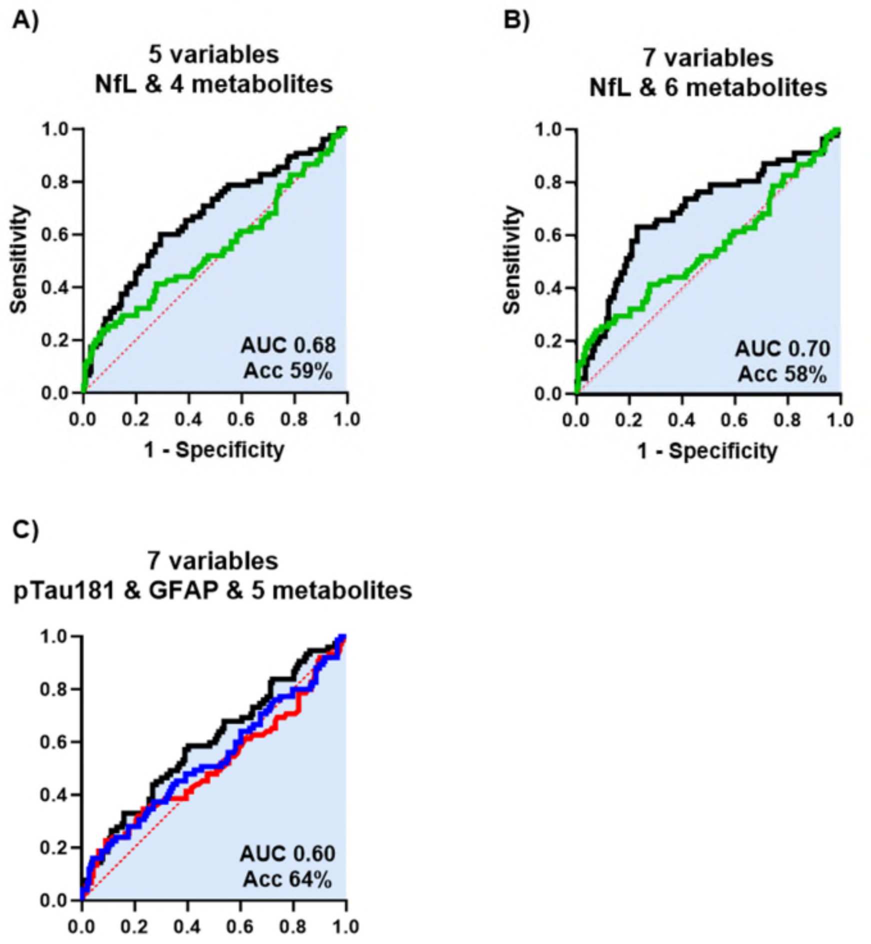


**SI Figure 6. Cross-validated ROC analysis of models prognosticating brain atrophy in UK Biobank. (A)** A five-variable model (NfL plus four metabolites: VLDL cholesterol, tyrosine, triglycerides, LDL particle diameter) achieved an AUC of 0.68 with 59% accuracy. **(B)** A seven-variable model (NfL plus six metabolites: VLDL cholesterol, triglycerides, valine, tyrosine, glutamine, LDL particle diameter) achieved an AUC of 0.70 with 58% accuracy. **(C)** A seven-variable model (pTau181 (red) and GFAP (blue) plus five metabolites: VLDL cholesterol, valine, tyrosine, acetate, alanine) achieved an AUC of 0.60 with 64% accuracy

**Validation of metabolomics results in the OPTIMA cohort**

To assess reproducibility, two seven-variable VITACOG metabolomics models were reconstructed using OPTIMA serum data. The first model included butyrylcarnitine, 2-hydroxyhexanoic acid, glucuronic acid, glutaric acid, histidine, and glutamate, while the second comprised butyrylcarnitine, 2-hydroxyhexanoic acid, glucuronic acid, glutaric acid, creatinine, and the NMR spectral region zgpr 3.40-3.68 ppm. When assessed using only the metabolomic features, excluding the protein biomarker pTau181, the respective AUCs in the VITACOG cohort were 0.85 and 0.86, compared with 0.88 and 0.91 for the corresponding multi-omic models. This indicates that the metabolomic signature alone retains strong discriminative ability.

Replication analyses in the independent OPTIMA cohort, comprising *post-mortem*-confirmed AD cases with baseline samples corresponding to an MCI-like cognitive range (MMSE 24-30), yielded AUCs of 0.81 and 0.80, comparable to those observed in the original VITACOG study (SI Fig. 7). The inclusion of neuropathologically verified AD provides a major advantage, confirming that the metabolomic signatures identified in VITACOG extend beyond prodromal stages to biologically defined disease. These findings underscore the reproducibility, robustness, and discriminatory power of the identified metabolic biomarkers for prognosticating cognitive decline in older adults with MCI-level impairment.

To further assess generalisability across neuropathological spectra, the same six-metabolite panels were evaluated in a broader OPTIMA subset encompassing mixed dementia cases, including both AD and vascular dementia (VaD) ($n$=61). Despite greater heterogeneity, both models retained good discriminative performance (AUCs 0.75 and 0.72; SI Fig. 8), indicating that these metabolic profiles capture cognitive decline–related processes beyond pure AD pathology. Although classification accuracy was lower than in neuropathologically confirmed AD, the preserved prognostic signal underscores the robustness of the identified metabolites and their relevance for progression in clinically and pathologically mixed MCI presentations. Together, the pure AD and mixed-pathology validations demonstrate that the metabolomic signatures derived from VITACOG are reproducible, biologically grounded, and informative of cognitive decline across diverse aetiological backgrounds.

**Validation of metabolomics results in the OPTIMA cohort – pure AD diagnosis**

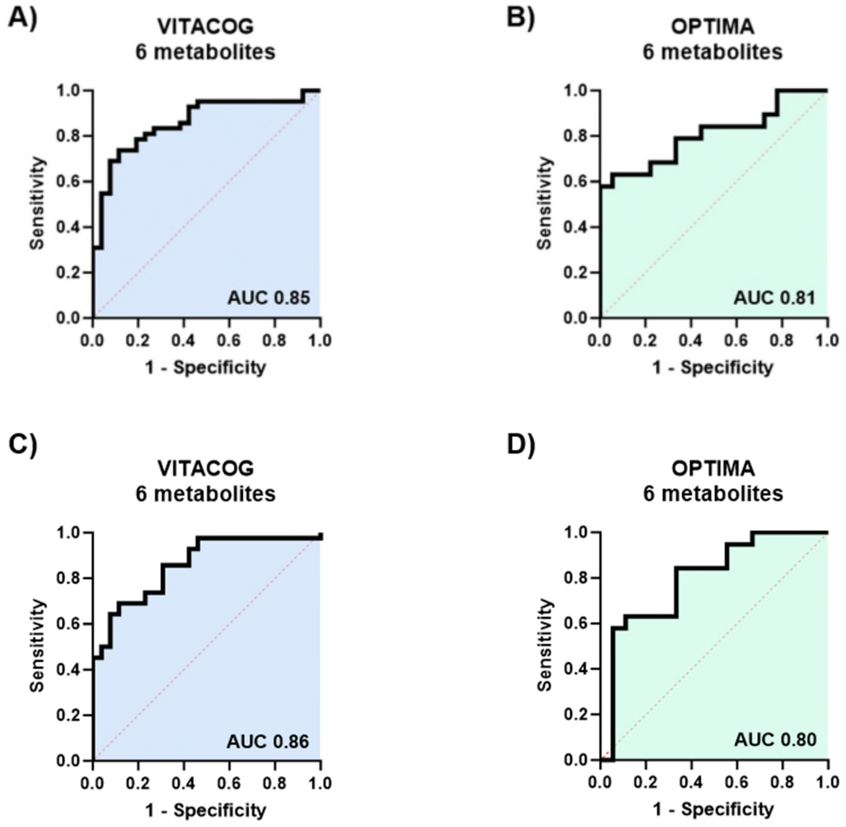


**SI Figure 7. Validation of VITACOG metabolomics models in the OPTIMA cohort.** Receiver operating characteristic (ROC) curves showing prognostic performance of multi-metabolite models for distinguishing progressors from stable individuals. **(A, B)** Model 1 comprising butyrylcarnitine, 2-hydroxyhexanoic acid, glucuronic acid, glutaric acid, creatinine, and the NMR spectral region zgpr30 3.40-3.68 ppm achieved an AUC of 0.85 in the VITACOG discovery cohort **(A)** and 0.81 in the OPTIMA validation cohort **(B)**. **(C, D)** Model 2 comprising butyrylcarnitine, 2-hydroxyhexanoic acid, glucuronic acid, glutaric acid, histidine, and glutamate achieved an AUC of 0.86 in VITACOG **(C)** and 0.80 in OPTIMA **(D)**. Both models retained good discriminatory performance in the independent OPTIMA cohort, confirming reproducibility of metabolite-based signatures for prognosticating cognitive decline in ageing individuals with mild cognitive impairment-level impairment.

**Validation of metabolomics results in the OPTIMA cohort – mixed diagnosis**

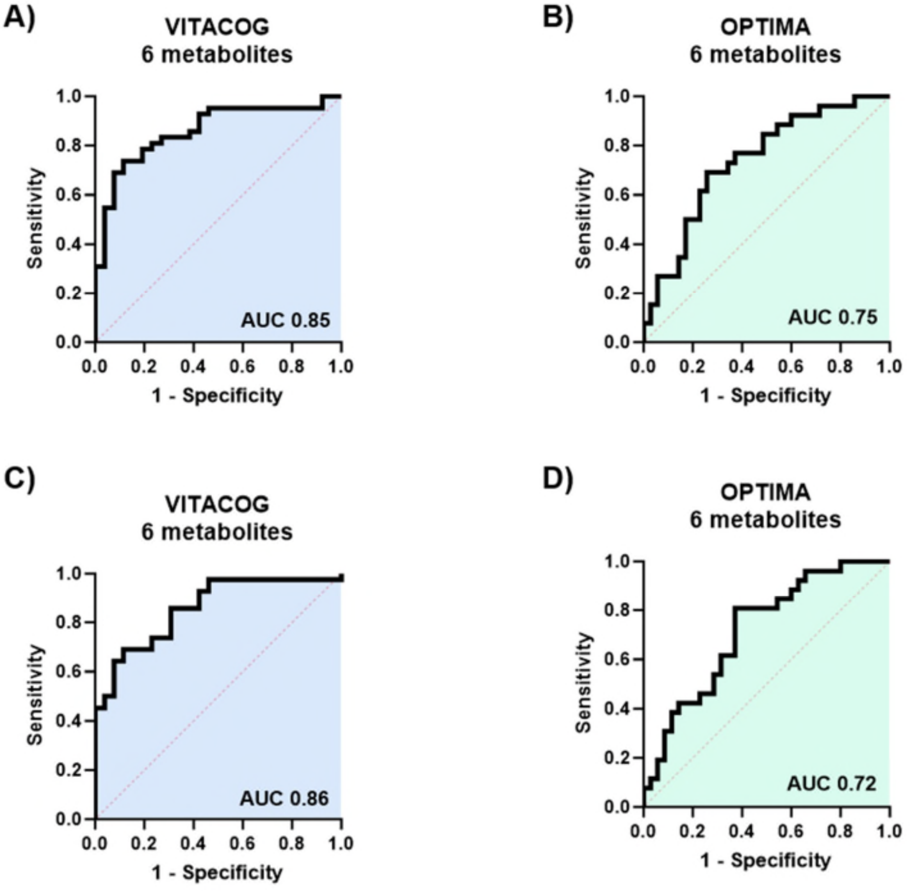


**SI Figure 8. Validation of VITACOG metabolomics models in the mixed-pathology OPTIMA cohort.** Receiver operating characteristic (ROC) curves showing prognostic performance of multi-metabolite models for distinguishing progressors from stable individuals in the mixed-pathology subset of OPTIMA (including AD and vascular dementia (VaD)). **(A, B)** Model 1 comprising butyrylcarnitine, 2-hydroxyhexanoic acid, glucuronic acid, glutaric acid, creatinine, and the NMR spectral region zgpr30 3.40–3.68 ppm achieved an AUC of 0.85 in the VITACOG discovery cohort **(A)** and 0.75 in the OPTIMA validation cohort **(B)**. **(C, D)** Model 2 comprising butyrylcarnitine, 2-hydroxyhexanoic acid, glucuronic acid, glutaric acid, histidine, and glutamate achieved an AUC of 0.86 in VITACOG **(C)** and 0.72 in OPTIMA **(D)**. Despite greater heterogeneity, both models maintained good discriminatory performance, demonstrating that the metabolite-based signatures identified in VITACOG remain informative of cognitive decline across clinically and pathologically mixed MCI presentations.

**Incorporation of targeted tHcy measurement reveals complementary predictors and enhances multi-omics model accuracy**

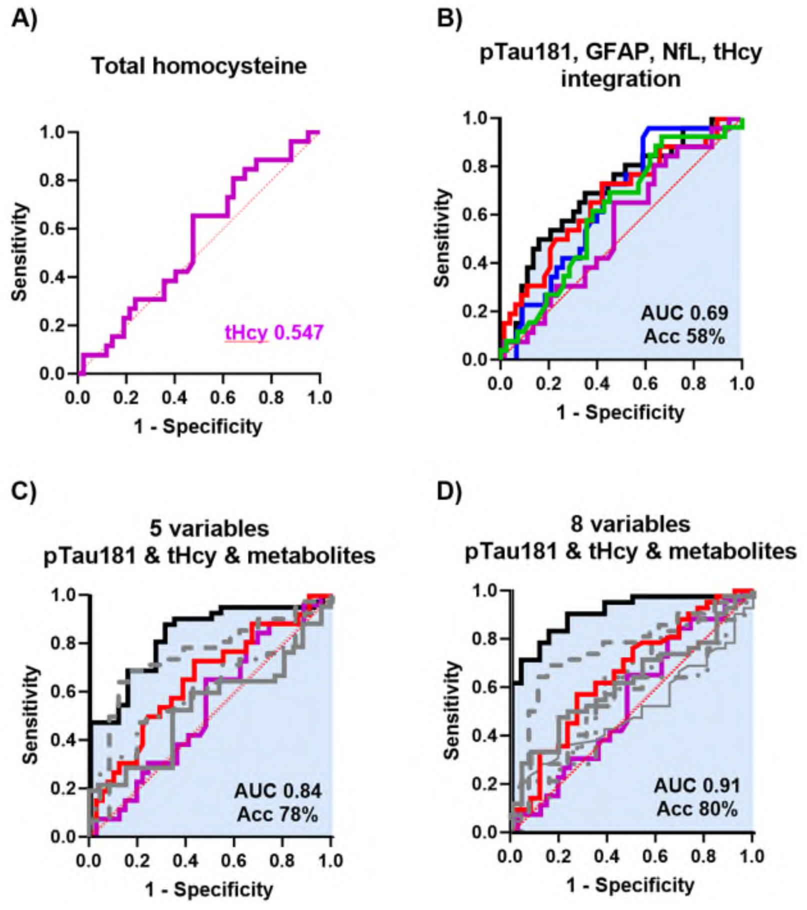


**SI Figure 9. Impact of targeted total homocysteine (tHcy) on multivariable model performance.** ***(A)*** tHcy alone showed limited discriminatory ability (AUC 0.547, pink). ***(B)*** Integration of tHcy with pTau181, GFAP, and NfL improved classification accuracy to 58% (tHcy pink, pTau181 red, NfL green, GFAP blue). ***(C)*** A five-variable model including pTau181, tHcy, and LC-MS/NMR metabolites achieved an AUC of 0.84 and accuracy of 78%, comparable to top untargeted metabolite-based models (tHcy pink, ptau181 red, glutaric acid grey, hydroxyhexanoic acid grey dashed line, butyrylcarnitine grey dash-dot line). **(D)** An eight-variable model combining pTau181 (red solid line), tHcy (pink solid line), glucuronic acid (AEC-MS, dash-dot line), 2-hydroxyhexanoic acid (AEC-MS, dashed line), butyrylcarnitine (RPLC-MS, bold long-dash line), histidine (CPMG, dotted line), glutaric acid (AEC-MS, triple-dot-dash line), and glutamine (CPMG, thin long-dash line) achieved an AUC of 0.91 and an accuracy of 80%. This model replicates that shown in main text Figure 4B, with the addition of tHcy, which did not improve prognostic performance.
